# HEPA Air Filters for Preventing Wildfire-Related Asthma Complications, a Cost-effectiveness Study

**DOI:** 10.1101/2023.04.17.23288697

**Authors:** Amin Adibi, Prabjit Barn, Erin M Shellington, Stephanie Harvard, Kate M Johnson, Christopher Carlsten

## Abstract

**Rationale:** Air pollution caused by wildfire smoke is linked to adverse health outcomes, especially for people living with asthma. We studied whether government rebates for high-efficiency particulate air (HEPA) filters, which reduce smoke particles indoors, are cost-effective in managing asthma and preventing exacerbations in British Columbia (BC), Canada.

**Methods:** A Markov model analyzed health states for asthma control, exacerbation severity, and death over a retrospective time-horizon of 5 years (2018-2022). Wildfire smoke-derived particulate matter (PM_2.5_) from the CanOSSEM model and relevant literature informed the model. The base case analysis assumed continuous use of the HEPA filter. Costs and quality-adjusted life-years (QALYs) resulting from varying rebates were computed for each Health Service Delivery Area (HSDA).

**Results:** In the base case analysis, HEPA air filter use resulted in increased costs of $83.34 (SE=1.03) and increased QALYs of 0.0011 (SE=0.0001) per person. Average incremental cost effectiveness ratio (ICER) among BC HSDAs was $74,652/QALY (SE=3,517), with ICERs ranging from $40,509 to $89,206 per QALY in HSDAs. Across the province, the intervention was projected to prevent 4,418 exacerbations requiring systemic corticosteroids, 643 emergency department visits, and 425 hospitalizations during the 5-year time horizon. A full rebate was cost-effective in one of the 16 HSDAs across BC. The probability of cost-effectiveness ranged from 0.1% to 74.8% across HSDAs. A $100 rebate was cost-effective in most HSDAs.

**Conclusions:** Our results indicate variable cost-effectiveness of HEPA filters in managing wildfire smoke-related asthma issues in BC. The effectiveness of government rebates varies by region but rebates up to two-thirds of the filter cost generally appear cost-effective, with a full rebate only cost-effective in Kootenay Boundary.

**Lay Summary:** Wildfire smoke can increase flare ups of symptoms among people living with asthma. These flare ups may require a visit to the emergency department or hospital admission. Research shows that portable HEPA air filters can significantly reduce concentrations of fine particles (PM2.5, an important component of wildfire smoke) in homes and other buildings. Using air filters during smoke events is a common public health recommendation. However, air filters are not accessible to everyone, with units costing anywhere between $150 to a few hundred dollars. Does it make sense for the government of BC to offer a rebate on the cost of purchasing air filters for every person living with asthma in BC? In this study, we used historical data on wildfire smoke concentrations between 2018 to 2022, computer simulations, and health economics methods to answer this question. Our results suggest that it is likely cost-effective for the government to pay for a portion of the costs of air filters, particularly in the interior and northern interior parts of BC. We also looked at other scenarios, such as filter use only when outdoor pollution exceeds certain thresholds that typically trigger an air quality advisory. We found that a $100 rebate was cost-effective when the air filter was used continuously, whereas a $30 rebate was cost-effective when the air filter was turned on only during air quality advisories.

## Background

The number, size, and intensity of wildfires in Canada have increased, particularly in the western province of British Columbia (BC) with the number of days with uncontrolled wildfire in BC expected to double or triple by 2100 (1).

Wildfire smoke is composed of several pollutants, including fine particulate matter with diameters of 2.5 microns and smaller (PM_2.5_). PM_2.5_ and other air pollutants have been associated with increased respiratory symptoms, hospitalizations, and other adverse health effects in individuals with asthma (2).

People living with asthma are particularly susceptible to air pollution. Asthma exacerbations (also known as *flare-ups* or *acute severe asthma*) are episodes characterized by progressive worsening of cough, wheezing, shortness of breath and decrease in lung function (3). Severe exacerbations can be fatal and can occur even in patients with well-controlled asthma (3). Previous studies have shown that PM_2.5_ from wildfire events can increase the risk of asthma exacerbations (4).

During wildfire smoke events, indoor PM_2.5_ concentrations increase as smoke infiltrates into homes and other buildings. Because people typically spend >70% of their time in indoor environments, (5) indoor air quality is an important contributor to total air pollution exposure. Consequently, interventions that improve indoor air quality are important to protecting health, particularly during episodes of poor air quality, such as during wildfire smoke events. Portable high-efficiency particulate air (HEPA) filters can reduce indoor concentrations of PM_2.5_(6). These units work by drawing air across a highly efficient filter that traps particles, including PM_2.5_, and release filtered air. HEPA and other filters can be portable, or be part of a building’s heating, ventilation, and air conditioning (HVAC) system, which is often referred to as induct filters.

As the climate emergency worsens and continues to impact air quality (7), there is growing consensus among the public health community that using air filters in indoor settings is an important health-protective intervention, particularly for vulnerable people. The Government of Canada currently provides tax benefits for the full cost of an air filter, cleaner, or purifier and up to $1000 for the purchase of an air conditioner for patients living with a chronic disease who have a prescription for these devices (8). In 2021, the Canadian Government also announced a 25% tax credit for small businesses to upgrade their ventilation systems and purchase portable HEPA air filters (9). In BC, the First Nations Health Authority (FNHA) provides portable air filters to communities affected by wildfires (10). We are also aware of two similar programs in the US: an air filter distribution program for low-income asthma patients by the Bay Area Air Quality and Management District (11), and a HEPA filter loaner program by the Forest Stewards Guild in Santa Fe (12). However, we are not aware of any formal analysis evaluating the cost-effectiveness of these programs from a health economics perspective.

In this study, we used a decision-analytic model to evaluate the cost-effectiveness of a government-sponsored portable HEPA air filters rebate program for improving asthma control and preventing asthma exacerbations caused by wildfire events in BC, Canada. Our analysis can serve as a blueprint for evaluating similar climate change adaptation strategies in BC and elsewhere. Some of the results of this study have been previously reported in the form of two abstracts (13, 14).

## Methods

We have reported the results of this study according to the recommendations and best practices set forth in the Consolidated Health Economic Evaluation Reporting Standards 2022 (CHEERS 2022) statement (15).

Based on discussions with policy makers and knowledge users in BC, we chose Health Service Delivery Area (HSDA) as the geographical unit of analysis. Our base case analysis assumes that the provincial government will offer a 100% rebate for portable HEPA air filters to all individuals diagnosed with asthma in BC. We used a retrospective time horizon of five years beginning in 2018 to the end of 2022, which was the most recent 5-year time horizon for which the data were available. This retrospective time horizon was necessary as daily projections of future wildfire PM_2.5_ concentrations are not available. We assumed patients on average spent 69.6% of their time at home (and thus could benefit from the air filter for the proportion of time they were at home), based on the time use information collected in Statistics Canada’s General Social Survey (5). The target population was BC residents diagnosed with asthma with a starting age of 42 (the average age of BC residents) (16).

We projected costs in 2023 Canadian dollars and effects as Quality-Adjusted Life Years (QALYs) for patients with and without portable HEPA air filters in their homes, and report results for each Health Service Delivery Area (HSDA) in BC. We also report the number of averted cases of asthma exacerbations using model-projected exacerbation rates and crude asthma prevalence levels from April 1st, 2020, to March 31st, 2021, for each HSDA obtained from BCCDC (17). We calculated Incremental Cost-effectiveness Ratios (ICER) and Net-Monetary Benefit (NMB) and reported cost-effectiveness at willingness-to-pay (WTP) threshold of $50,000/QALY.

The analysis was conducted from the healthcare payer perspective, with an annual discounting of 1.5% applied to costs and effects.

### Stakeholder engagement

We developed a health economic analysis plan with early and ongoing input from stakeholders, including two patient partners living with asthma, two medical health officers, an environment health officer, and a policy analyst (see Acknowledgment section).

### Model development

We developed a time-varying Markov model with seven health states corresponding to well-controlled asthma, partly-controlled asthma, and uncontrolled asthma (as defined per Global Initiative for Asthma (GINA) 2023 (3)) as well as exacerbations requiring either systemic corticosteroids (ExacSCS), a visit to the Emergency Room (ExacER), or hospitalization (ExacHosp), and death, respectively (Figure 1).

**Figure 1:**
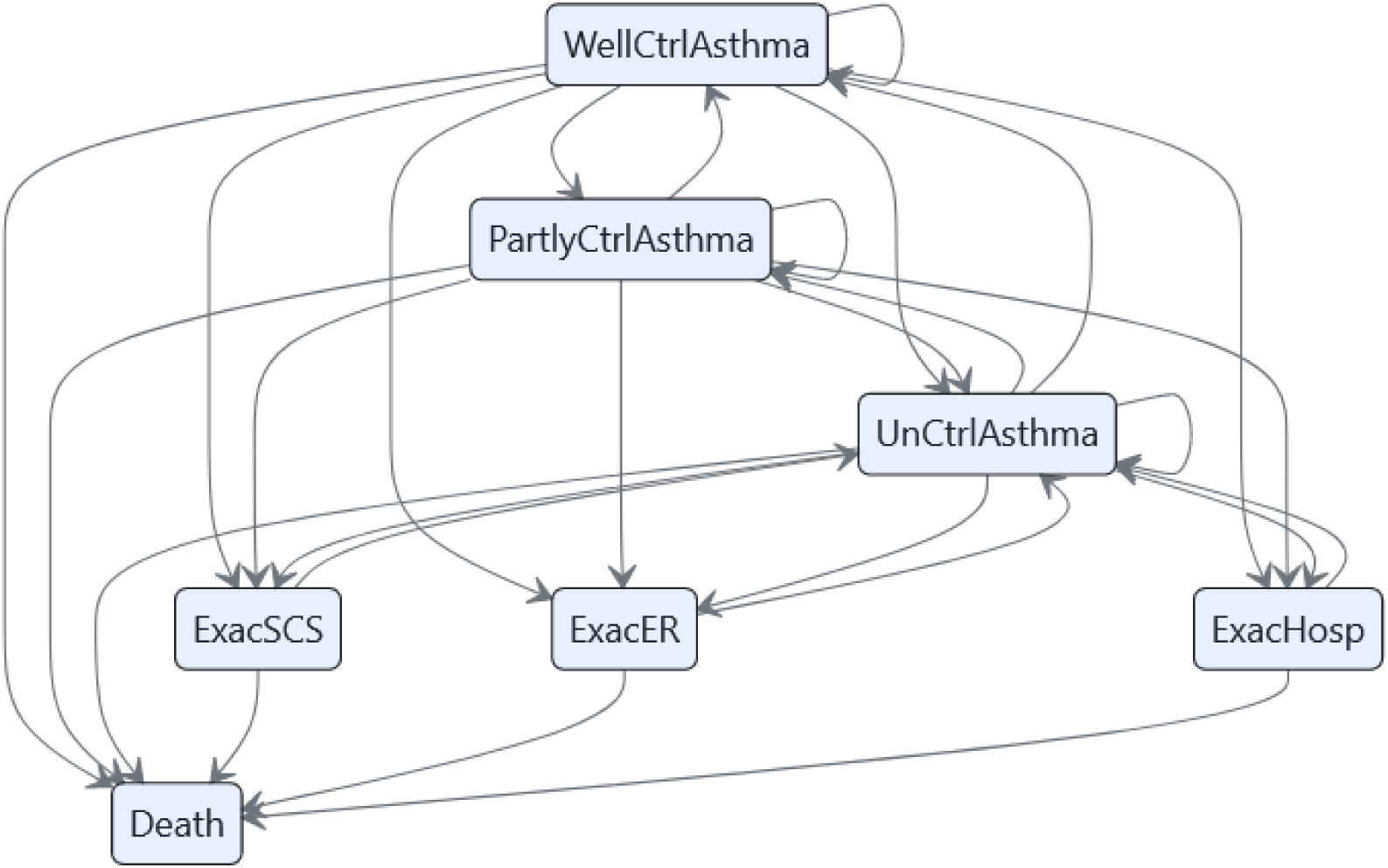
Markov health states and transitions. WellCtrlAsthma = Well-controlled asthma; UnCtrlAsthma = Uncontrolled Asthma; Exac-SCS = Exacerbations requiring systemic corticosteroids; ExacER = Exacerbation requiring a visit to the emergency room; ExacHosp = Exacerbations requiring hospitalization.

Background mortality was based on age-specific life tables for BC from Statistics Canada (18). Mortality due to asthma exacerbations of each severity was based on a national review of asthma deaths in the UK (19, 20). Annual transition probabilities between asthma control states were based on an original analysis of Economic Burden of Asthma study where we calculated the proportion of transitions occurring between each control state over 5 visits conducted over 1 year of follow-up (21). Rates of severe exacerbations leading to SCS, ER, or hospitalization were obtained from SYGMA II study (22). We applied a risk ratio of 1.40 to individuals with partially controlled and uncontrolled asthma to reflect their higher probability of exacerbation. This parameter was based on an analysis of commercially insured patients in the US (23).

We ran the model using daily time cycles.

### Air Pollution Exposure

Average daily outdoor PM_2.5_ concentrations were obtained from CanOSSEM (24), a random forest machine learning model developed and validated by BC Center for Disease Control that projects retrospective average daily wildfire smoke levels for each postal code in BC. Outdoor PM_2.5_ concentrations in HSDAs were obtained by linking postal codes to HSDAs using Postal Code Conversion File Plus (PCCF+) Version 7E (25). Model assumptions are listed in Table 1.

**Table 1:**
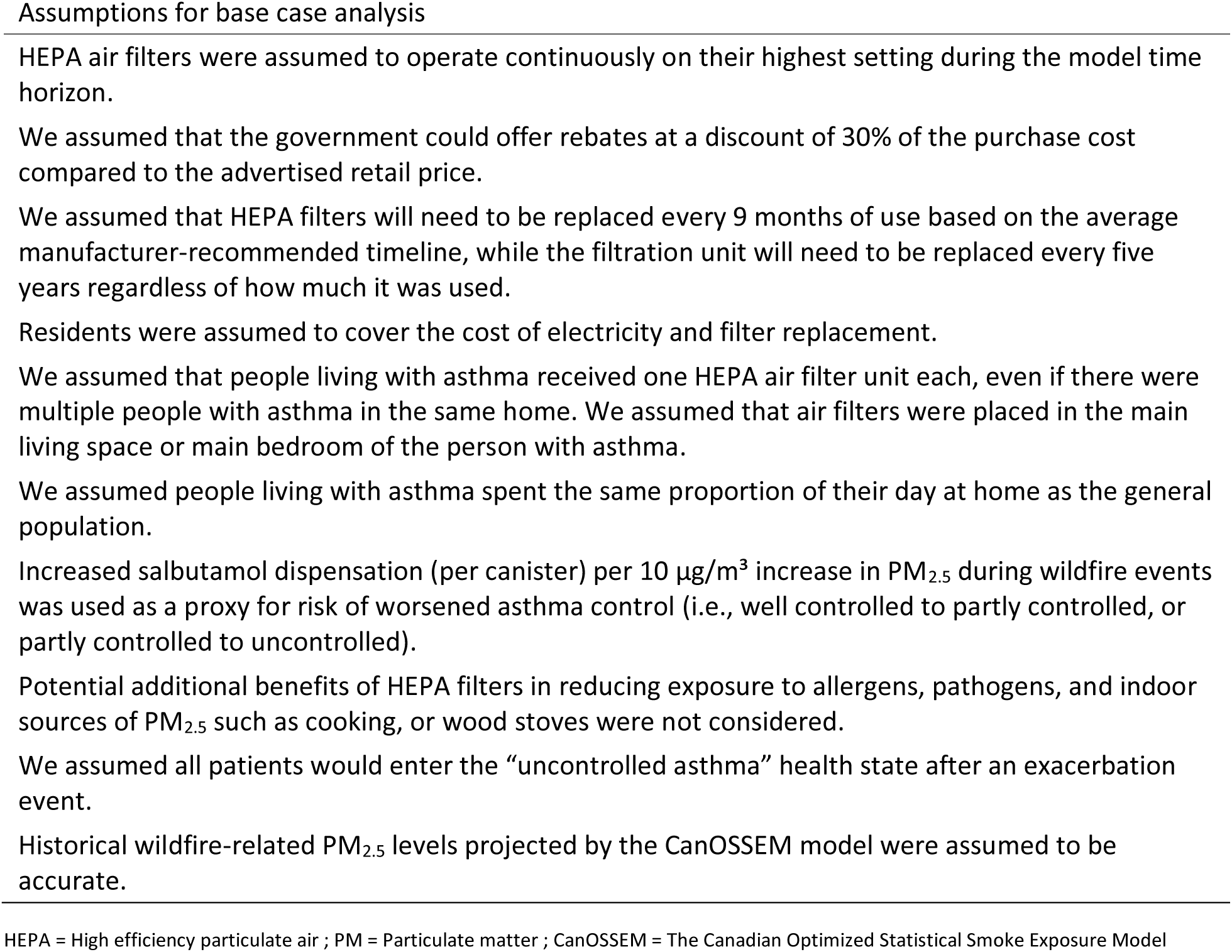
Model assumptions for evaluating cost-effectiveness of a portable air filter rebate program to prevent asthma exacerbations.

| Assumptions for base case analysis |
| --- |
| HEPA air filters were assumed to operate continuously on their highest setting during the model time horizon. |
| We assumed that the government could offer rebates at a discount of 30% of the purchase cost compared to the advertised retail price. |
| We assumed that HEPA filters will need to be replaced every 9 months of use based on the average manufacturer-recommended timeline, while the filtration unit will need to be replaced every five years regardless of how much it was used. |
| Residents were assumed to cover the cost of electricity and filter replacement. |
| We assumed that people living with asthma received one HEPA air filter unit each, even if there were multiple people with asthma in the same home. We assumed that air filters were placed in the main living space or main bedroom of the person with asthma. |
| We assumed people living with asthma spent the same proportion of their day at home as the general population. |
| Increased salbutamol dispensation (per canister) per 10 $\mu\text{g}/\text{m}^3$ increase in $\text{PM}_{2.5}$ during wildfire events was used as a proxy for risk of worsened asthma control (i.e., well controlled to partly controlled, or partly controlled to uncontrolled). |
| Potential additional benefits of HEPA filters in reducing exposure to allergens, pathogens, and indoor sources of $\text{PM}_{2.5}$ such as cooking, or wood stoves were not considered. |
| We assumed all patients would enter the “uncontrolled asthma” health state after an exacerbation event. |
| Historical wildfire-related $\text{PM}_{2.5}$ levels projected by the CanOSSEM model were assumed to be accurate. |

HEPA = High efficiency particulate air ; PM = Particulate matter ; CanOSSEM = The Canadian Optimized Statistical Smoke Exposure Model

Risk ratios for the effect of increased exposure to PM_2.5_ on asthma outcomes, including salbutamol dispensation and asthma-related physician visit, ER visit, and hospitalizations were obtained from a recent meta-analysis (4) and a model validation study based on BC administrative health data (26).

Transition probabilities, utility and disutility values, rate ratios for the effect of increased PM_2.5_ pollution on asthma outcomes, healthcare state costs, outdoor to indoor PM_2.5_ infiltration rates, and HEPA filter efficiency rate were obtained from the literature (Table 2).

**Table 2:**
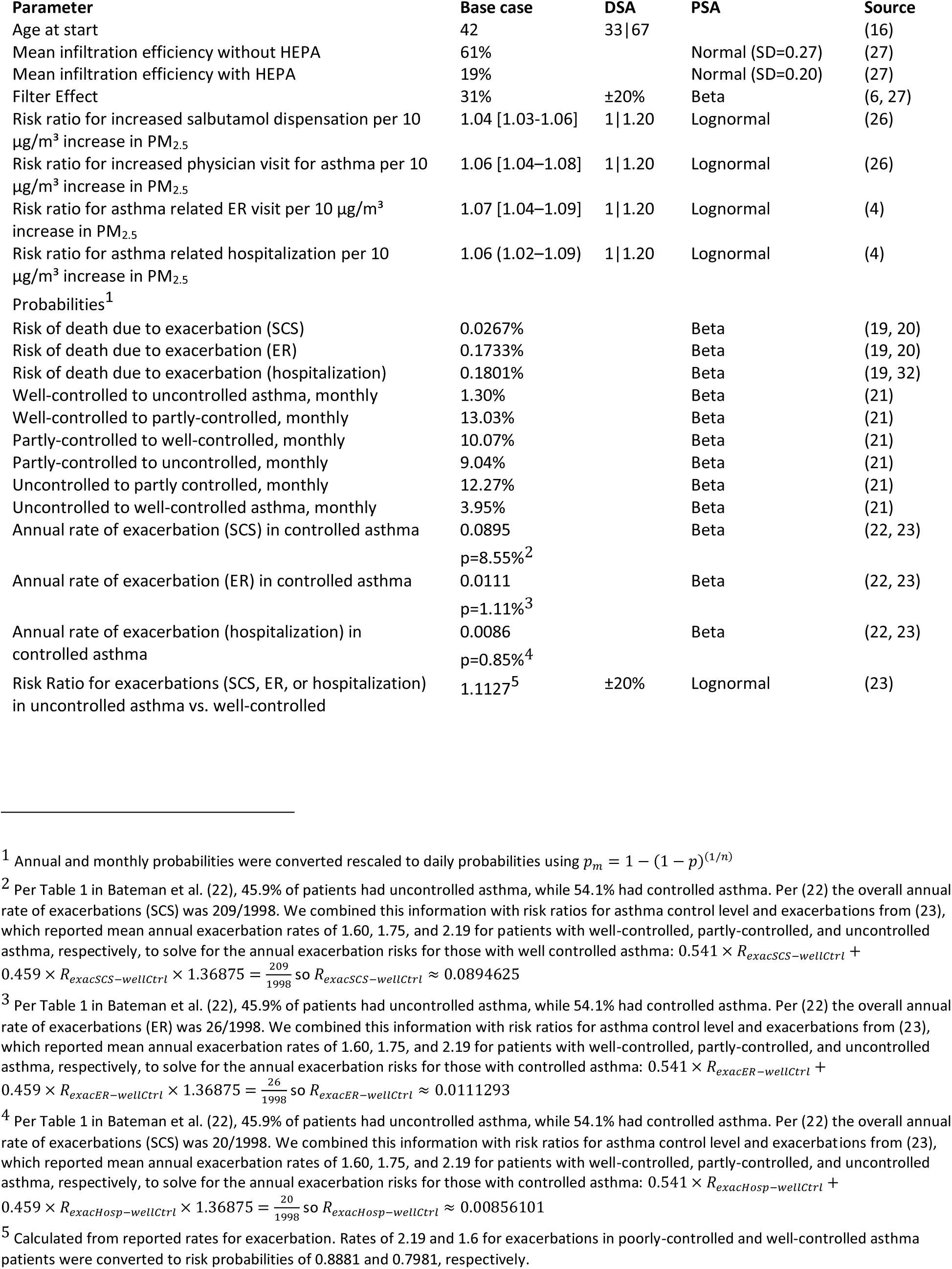
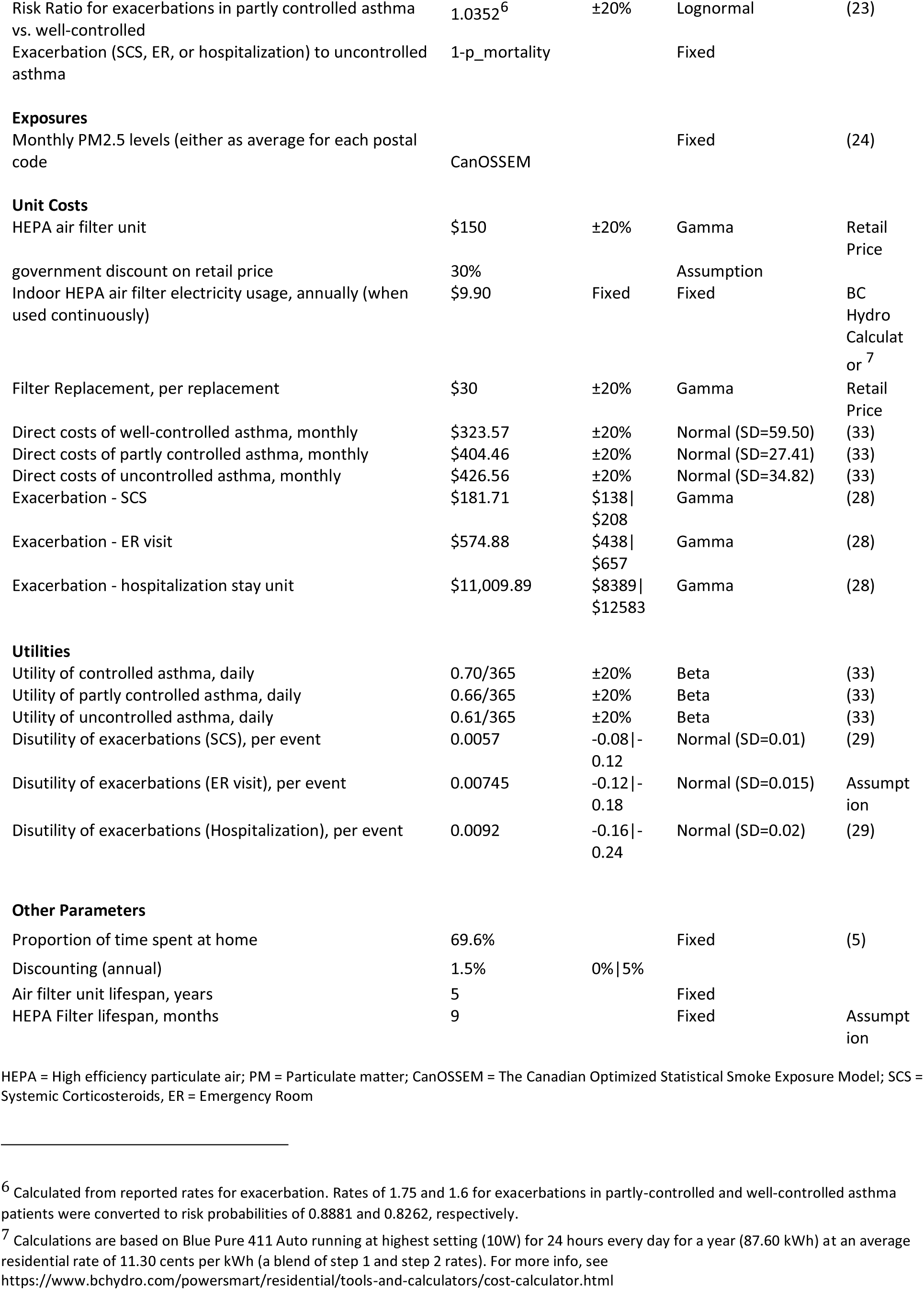
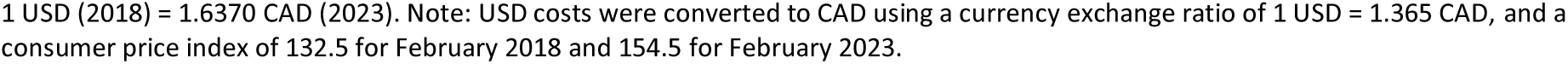
Model Parameters.

### HEPA Air Filter Effectiveness

We chose what we considered to be a *typical* HEPA air filter unit with a clean air delivery rate (CADR) of 105 cfm for smoke, and a nominal air exchange rate of 4.8/hr for a coverage area of 15 m^2^. Measured HEPA filter efficiency of 0.31 (defined as the ratio of indoor PM_2.5_ measured throughout the year with HEPA to without HEPA filter) was obtained from a study led by one of our co-authors that evaluated air filter effectiveness in BC homes during smoke events (27) using a comparable air filter unit with a CADR of 150 cfm and nominal air exchange rate of 6/hr for a coverage area of 17.37 m^2^ (Table 2). Varying filter effectiveness values of (±20%) were explored in one-way sensitivity analysis.

### Costs

Costs included the initial purchase price of the HEPA air filter unit, background healthcare costs based on asthma control level, and unit costs of exacerbations obtained from the literature. Unit costs and utilization were obtained from previous studies (22, 28).

Costs to patients for air filter operation such as electricity and replacement HEPA filters after every 9 months of use (based average replacement duration according to the manufacturer) were not included in the base case analysis but are reported in scenario analysis.

### Health State Utilities

Health state utilities were derived from the literature based on levels of asthma control, while severe exacerbations requiring systemic SCS, ER visit, or hospitalization were associated with a one-time disutility value derived from EQ-5D questionnaires (29).

### Sensitivity Analyses

One-way deterministic sensitivity analysis was used to explore the effect of changing assumptions on the estimated costs and QALYs. Uncertainty in the results due to parameter uncertainty was explored through probabilistic sensitivity analysis with 1000 sampling from parameters distributions (Table 2) in each HSDA.

Our base case scenario assumed that the government covered the full cost of the air filter and that air filters were operating continuously throughout the five years of study. Here, we explore three different scenarios: 1) the government pays a $100 (67%) rebate, 2) the government pays a full (100%) rebate and air filters are turned on only when the outdoor pollution exceeds certain thresholds, and 3) the government pays a $30 (20%) rebate, and the air filter operates only when outdoor PM_2.5_ concentration is above a certain threshold. We chose rebate amounts based on convenience and existing provincial rebate programs (e.g., for energy efficient products (30).

### Software

Data preparation, model development, and statistical analysis was performed in R v4.3.1 using the *heemod* package v0.15.1 (31). We used Quarto v1.4.346 to create a reproducible manuscript and used version control to keep track of methodological decisions and changes to the model. Model code is publicly available at https://github.com/resplab/hepa_wildfire_CE_code.

## Results

Average daily wildfire-related smoke concentration ranged from 2.5 μg/m^3^ (2019-09-25, Northeast) to 410.6 μg/m^3^ (2018-08-19, Kootenay Boundary). Significant year-to-year variability was observed among all HSDAs, with higher smoke concentration during years with more wildfire activity in the Interior and Northern Health regions as shown in Figure 2.

**Figure 2:**
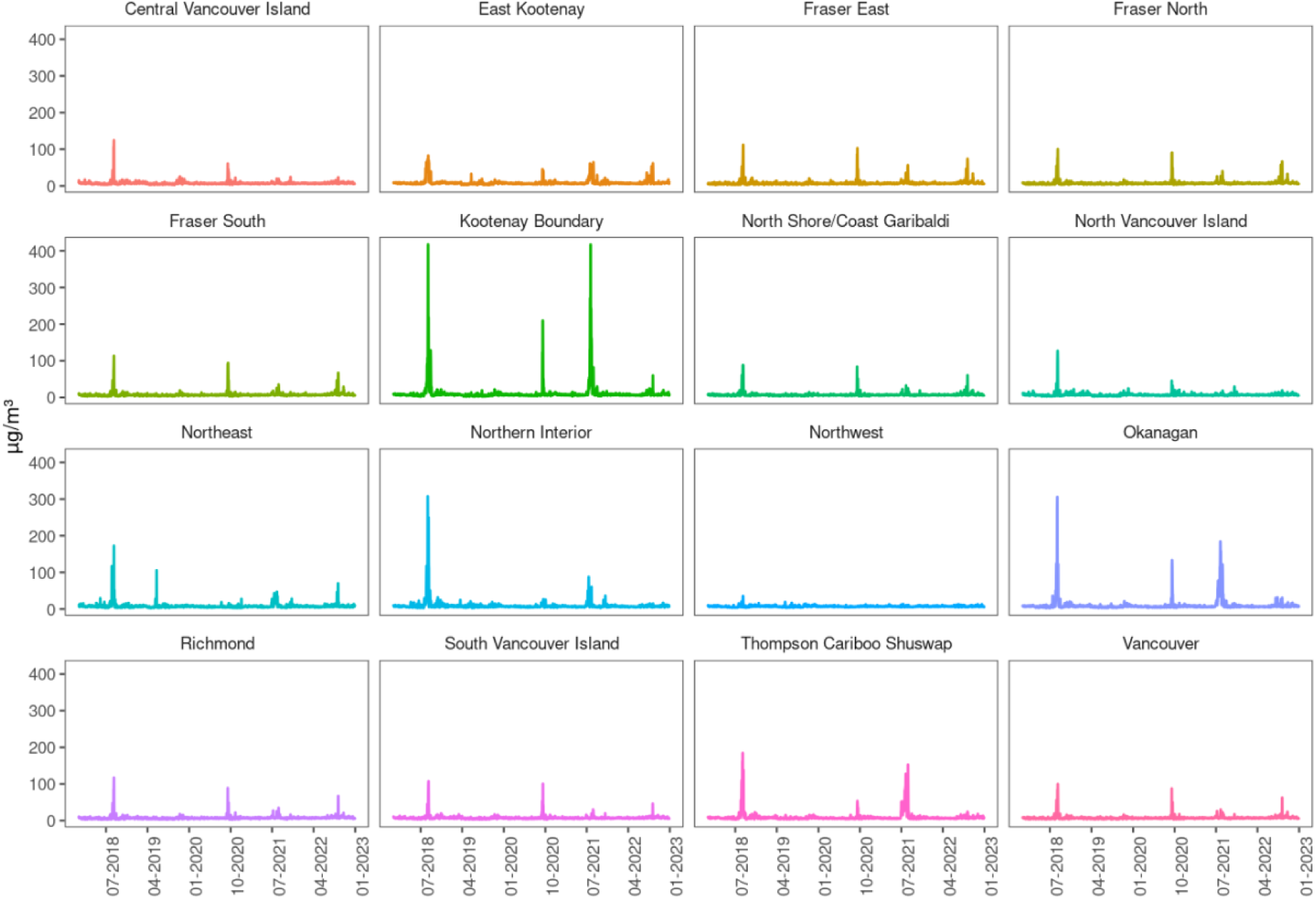
Daily Smoke Exposure Levels (PM2.5) across BC

### Base Case Cost-Effectiveness

Figure 3 shows the incremental cost-effectiveness ratio (ICER) for each HSDA in BC during the time horizon and the associated probability of cost-effectiveness when the uncertainty around model input parameters (Table 2) is taken into account. In the base case analysis in which government paid 100% of the purchase cost for HEPA filter units, ICER was below a WTP threshold of $50,000/QALY in Kootenay Boundary and above the threshold elsewhere in the province.

**Figure 3:**
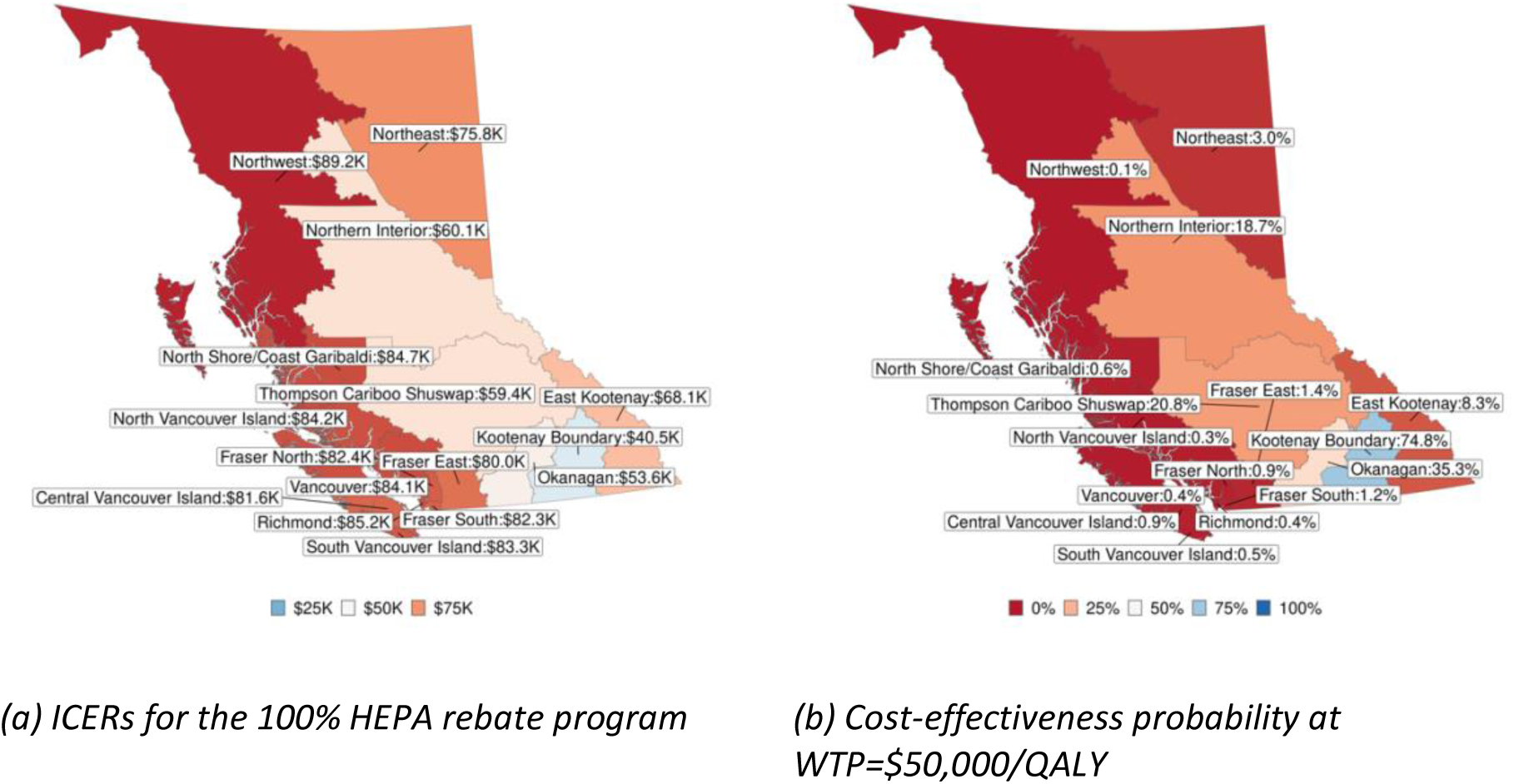
Base-case results

Table 3 ranks HSDAs in BC in terms of HEPA rebate program cost-effectiveness, in descending order based on ICER. ICERs ranged from $40,509/QALY in Kootenay Boundary to $89,206/QALY in Northwest. Based on model projections and prevalence of asthma in BC, a total of 4,418 severe exacerbations leading to systemic corticosteroids use, 643 emergency department visits, and 425 cases of hospitalizations could be averted by continuous HEPA air filter use. Due to the larger populations and higher prevalence of asthma, the highest number of severe exacerbations averted (including systemic corticosteroids use, emergency department visits, and hospitalizations) were in Fraser South (961), Fraser North (644), Okanagan (607), and Vancouver (590).

**Table 3:**
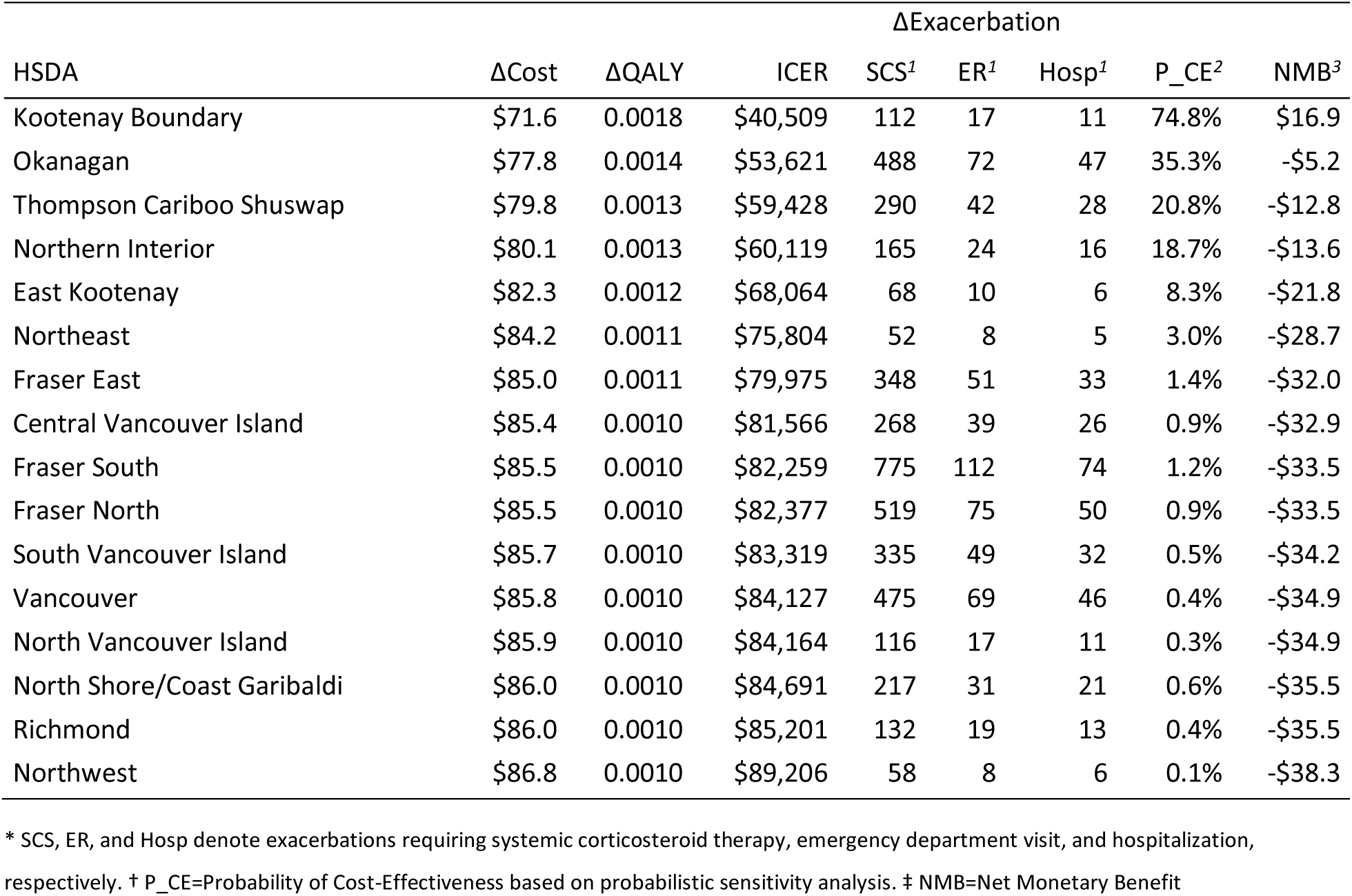
ICERs for the portable HEPA air cleaner rebate program in BC.

Cost-effectiveness probabilities were highest in Kootenay Boundary (74.8%), Okanagan (35.3%), and Thomson Cariboo Shuswap (20.8%) HSDAs. One-way sensitivity analysis (Appendix) showed that costs and QALYs were most sensitive to the risk ratios of increased salbutamol dispensation and hospitalization per 10 µg/m³ increase in PM_2.5_, utility of well-controlled and uncontrolled asthma, and the retail price of air filter units.

### Scenario Analyses

Figure 4 shows results of scenario analyses. Our results suggest that a $100 rebate program would have been cost-effective at a WTP threshold of $50,000/QALY everywhere in the province except for the North Shore/Coast Garibaldi and Northwest HSDAs with ICERS of $50,500/QALY and $53,200/QALY, respectively.

**Figure 4:**
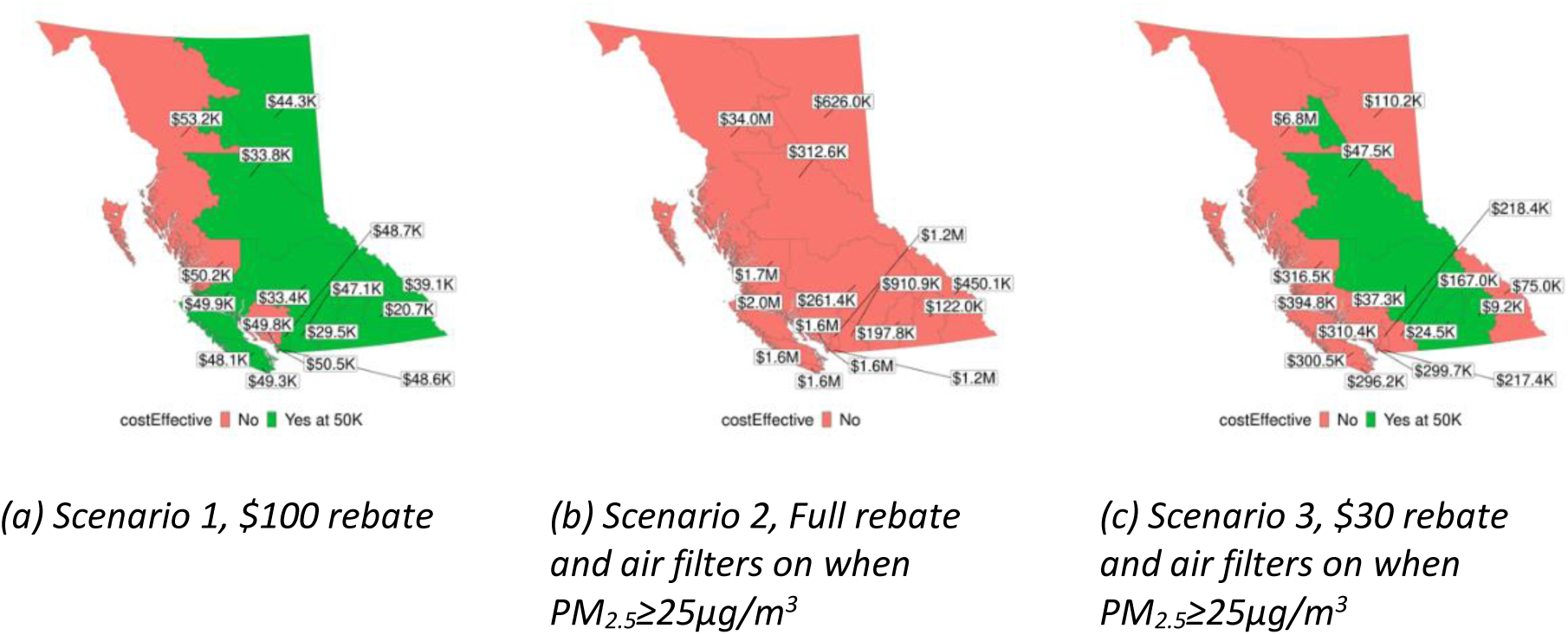
ICERs for HEPA Rebate Program - Different Scenarios

The next two scenarios are based on the operation of HEPA filters when the outdoor PM_2.5_ exceeded a threshold concentration. We used a threshold of 25 μg/m^3^ for PM_2.5_, based on the BC government 24-hour ambient air quality objective which is used, along with other information to guide decisions on when to issue an air quality advisory (34).

Days with PM_2.5_ concentrations above 25 μg/m^3^ were most common in August, followed by September, July, October, and May. Our results suggest that a full purchase rebate along with operation of air filters on days in which outdoor PM_2.5_ concentrations exceeded 25 μg/m^3^ would not have been cost-effective anywhere in BC.

The last scenario considered a combination of a $30 rebate and operation of air filters on days in which PM_2.5_ concentrations exceeded 25 μg/m^3^. Our results suggest that the intervention would have been cost-effective in Kootenay Boundary, Okanagan, Thompson Cariboo Shuswap, and Northern Interior.

Other possible scenarios and the effect of alternative inputs on the results can be explored further using a web app, available at https://resplab.shinyapps.io/hepa_wildfire_CE/

### Operation Costs

While a formal evaluation of the intervention from a societal perspective is beyond the scope of this work, operation costs for patients were calculated to provide additional context. In the base case analysis when the air filter is operating continuously at its highest setting, patients anywhere in BC can expect to pay an average of $10 for 87.60 kWh of electricity and $40 for HEPA filter replacements annually, for a total of $50 per year. In the threshold-based scenarios, operation costs would be much lower (between $0.03 to $1.91) and across different HSDAs as shown in the Appendix Table A1.

## Discussion

We found that across BC, offering a 100% rebate on HEPA air filters was cost-effective between 2018-2022 in Kootenay Boundary HSDA, which had been the most wildfire prone HSDA in that time frame. Our results suggest that a $100 rebate program was cost-effective in most of the province when air filters were used continuously throughout the year. When air filters are only operated on days in which PM_2.5_ levels exceed 25 μg/m^3^, a $30 rebate program was also cost-effective in wildfire-prone areas of the interior and northern interior BC. To the best of our knowledge, this is the first cost-effectiveness analysis of a government-sponsored HEPA air filter rebate program designed to prevent wildfire smoke-related asthma exacerbations and improve asthma control.

Particulate matter pollution is a major cause of health and economic burden in Canada. In its 2022 report on the health of Canadians in a changing climate, Health Canada classified fine particulate matter among the three major outdoor pollutants which are collectively responsible for 15,300 premature deaths in Canada annually, with an economic cost of $114 billion (35). There are growing calls for governments to better protect health, including by covering the cost of climate adaptation measures that protect the public. For example, the BC Coroner’s report on the 2021 heat dome in BC, which resulted in 619 deaths, recommended that the BC government increase accessibility of air conditioners for use during extreme events by allowing them to be provided as medical devices through existing provincial programs (36). In response to the Coroner’s Report, the BC Government launched a new initiative in June 2023 to provide 8000 publicly funded air conditioning units to low-income and medically vulnerable individuals (37). Heat events and smoke can occur together, and the current public health advice is to create or access cool environments with clean air. Our results suggest that a similar program should be implemented for HEPA filter air cleaners to mitigate the impacts of extreme wildfire events in HSDAs with recurrently high wildfire smoke exposure. Considering the equity implications of such programs, we believe that offering rebates for portable HEPA air filters can enhance equal access to healthier indoor environments. Such rebates could extend affordability to renters too, since presently available rebates primarily target homeowners.

We made several assumptions to develop our cost-effectiveness model. Where possible, we opted for assumptions that would minimize the chance of wrongly identifying the intervention as cost-effective. For instance, we narrowly focused on the short-term health benefits of HEPA filters in preventing acute asthma complications. However, chronic exposure to wildfire smoke may also be associated with increased risk of asthma incidence. Maintaining asthma control and preventing exacerbations is likely associated with improved long-term respiratory outcomes, which were not accounted for in our analysis. We only considered the benefits of air filters in reducing exposure to wildfire-related PM_2.5_. However, HEPA air filters reduce concentrations of PM_2.5_ from all sources, including traffic and industry, indoor sources, allergens, bacteria, and respiratory viruses such as flu and COVID-19. We also assumed that HEPA air filter units would last only five years, regardless of how much they were in use, while the HEPA filters had to be replaced every 9 months.

We also assumed the individuals with asthma spent the same proportion of time indoors as the general public. However, it is plausible that people living with asthma might increase their time indoors on days with high levels of wildfire pollution, thereby improving the cost-effectiveness of portable HEPA filters compared to what we have reported.

In our base case analysis, we assumed the air filter to be turned on continuously for the 5-year time-horizon of the model, which is in line with Health Canada’s guideline that asserts there is no threshold of exposure to PM_2.5_ at which negative health effects may not occur (38). Continuous operation of air filters also ensures further benefits from reducing exposure to indoor sources of PM_2.5_, allergens, and reduced transmission of respiratory infections.

There might be concerns about the practicality of running portable HEPA filters continuously. Previous studies have shown that adherence might be negatively impacted because of the machine’s noise and the perceived cold draft from the machines, especially during winter (39). Our study implicitly accounts for this, as we have relied on real-world experimental measurements of filter effect that were done in summer and winter across BC (27).

The CanOSSEM model provides estimates for PM_2.5_ in general, with improving accounting for wildfire smoke. Therefore, our results reflect the impact of HEPA filters on PM_2.5_ attributable to all sources, although in the Pacific Northwest, wildfires are the biggest contributor to PM_2.5_ (40–42) Our scenario analyses also showed that continuous operation of the HEPA air filter is more beneficial than turning it on and off daily based on the provincial 24-hour PM_2.5_ ambient air quality objective. It makes sense for the continuous operation to be the most cost-effective choice from the government’s perspective since there is more benefit to reap with no additional cost as the government is only paying for the upfront cost of a rebate.

Several limitations should be noted. First, the stochastic and hard-to-predict nature of wildfire events prevented us from conducting this analysis prospectively, as long-term prediction of wildfire events in BC with adequate spatial and temporal resolution are not available. Our retrospective results are still useful for future planning, as the frequency and intensity of wildfires in BC is expected to grow, and higher levels of exposure will make the intervention more cost-effective.

Second, retrospective wildfire-related PM_2.5_ concentrations used in this study are based on the results of the CanOSSEM model, and thus subject to limitations and uncertainties of that model.

Third, within the observed PM_2.5_ concentration range of 2.3 μg/m^3^ to 417.3 μg/m^3^, we have assumed a linear dose-response relationship for increased risk of change in asthma control and asthma exacerbations leading to either SCS, ER visit, or hospitalizations.

Lastly, due to a lack of data, we did not evaluate HEPA air filters in subgroups of the population based on sex, age, ethnicity, or social determinants of health, despite their established impact on the burden of the disease (43–45).

## Conclusion

Between 2018 and 2022, offering a 100% rebate on portable HEPA air filters was a cost-effective intervention to reduce short-term asthma complications due to wildfire smoke in Kootenay Boundary but not in other HSDAs in BC. Consumer rebates of up to $100 (about two-thirds of the cost of the air filter unit) were a cost-effective alternative in most of the province, especially the interior and northern interior parts of the province where wildfire exposure is higher.

## Data Availability

All data produced in the present study are available upon reasonable request to the authors

https://github.com/resplab/hepa_wildfire_CE_code

https://resplab.shinyapps.io/hepa_wildfire_CE/

## Acknowledgments

We would like to acknowledge that most of the activities of this research project were conducted on the traditional, ancestral, and unceded territory of the Musqueam people. We would like to thank our patient partners for sharing their feedback and insights with us throughout this study. We are also thankful to our knowledge user advisers Dr. Michael Schwandt (Medical Health Officer, Vancouver Coastal Health), Dr. Silvina Mema (Medical Health Officer, Interior Health), Paula Tait (Health & Resource Development Technical Adviser at Northern Health), and Jade Yehia (Environmental Health Policy Lead at BC Ministry of Health). We would like to express our gratitude Dr. Mohsen Sadatsafavi (University of British Columbia) and Dr. Zafar Zafari (University of Maryland) for their help with health economics methods. Lastly, we are very thankful to Dr. Sarah Henderson and Naman Paul (BC Centre for Disease Control) for sharing the results of the CanOSSEM model with us. This study was funded by Legacy for Airway Health.

### Appendix: Operation Costs

**Table A1:** Average annual operation cost for patients under different scenarios.

| **HSDA** | **Base case, Scenario 1** | **Scenarios 2 and 3** |
| --- | --- | --- |
| Central Vancouver Island | \$50.00 | \$0.36 |
| East Kootenay | \$50.00 | \$1.61 |
| Fraser East | \$50.00 | \$0.70 |
| Fraser North | \$50.00 | \$0.61 |
| Fraser South | \$50.00 | \$0.55 |
| Kootenay Boundary | \$50.00 | \$1.91 |
| North Shore/Coast Garibaldi | \$50.00 | \$0.36 |
| North Vancouver Island | \$50.00 | \$0.25 |
| Northeast | \$50.00 | \$1.13 |
| Northern Interior | \$50.00 | \$1.20 |
| Northwest | \$50.00 | \$0.03 |
| Okanagan | \$50.00 | \$1.83 |
| Richmond | \$50.00 | \$0.36 |
| South Vancouver Island | \$50.00 | \$0.36 |
| Thompson Cariboo Shuswap | \$50.00 | \$1.66 |
| Vancouver | \$50.00 | \$0.36 |

### Appendix: Deterministic Sensitivity Analysis

The following plots show the effect of changing input parameters on overall ICER for each HSDA in each year.

Please refer to the following table for variable names and descriptions:

*Variable names and descriptions for Deterministic Sensitivity Analysis*

| Name | Description |
| --- | --- |
| age_init | Age at the start of the model time horizon |
| r_discount | Annual discounting of costs and utilities |
| prop_time_at_home | Average proportion of time spent at home |
| filter_effect | The ratio of mean infiltration efficiency with HEPA to without HEPA |
| rr_salbutamol | Risk ratio for increased salbutamol dispensation per 10 $\mu\text{g}/\text{m}^3$ increase in $\text{PM}_{2.5}$ |
| rr_gp | Risk ratio for increased asthma-related physician visit per 10 $\mu\text{g}/\text{m}^3$ increase in $\text{PM}_{2.5}$ |
| rr_er | Risk ratio for asthma related ER visit per 10 $\mu\text{g}/\text{m}^3$ increase in $\text{PM}_{2.5}$ |
| rr_hosp | Risk ratio for asthma related hospitalization per 10 $\mu\text{g}/\text{m}^3$ increase in $\text{PM}_{2.5}$ |
| rr_sev_exac_unctrl | Risk ratio for severe exacerbations in uncontrolled asthma vs. well-controlled |
| rr_sev_exac_part_ctrl | Risk ratio for severe exacerbations in partly controlled asthma vs. well-controlled |
| c_asthma_well | Direct costs of well-controlled asthma, monthly |
| c_asthma_part_ctrl | Direct costs of partly controlled asthma, monthly |
| c_asthma_unctrl | Direct costs of uncontrolled asthma, monthly |
| c_exac_scs | Direct cost of an exacerbation leading to systemic corticosteroid use |
| c_exac_er | Direct cost of an exacerbation leading to emergency visit |
| c_exac_hosp | Direct cost of an exacerbation leading to hospitalization |
| c_hepa_unit_list_price | HEPA air filter unit list price |
| du_exac_scs | Disutility of exacerbations leading to systemic corticosteroid use, per event |
| du_exac_er | Disutility of exacerbations leading to emergency visit, per event |
| du_exac_hosp | Disutility of exacerbations leading to hospitalization, per event |
| u_asthma_well | Utility of controlled asthma, daily |
| u_asthma_part_ctrl | Utility of partly controlled asthma, daily |
| u_asthma_unctrl | Utility of uncontrolled asthma, daily |

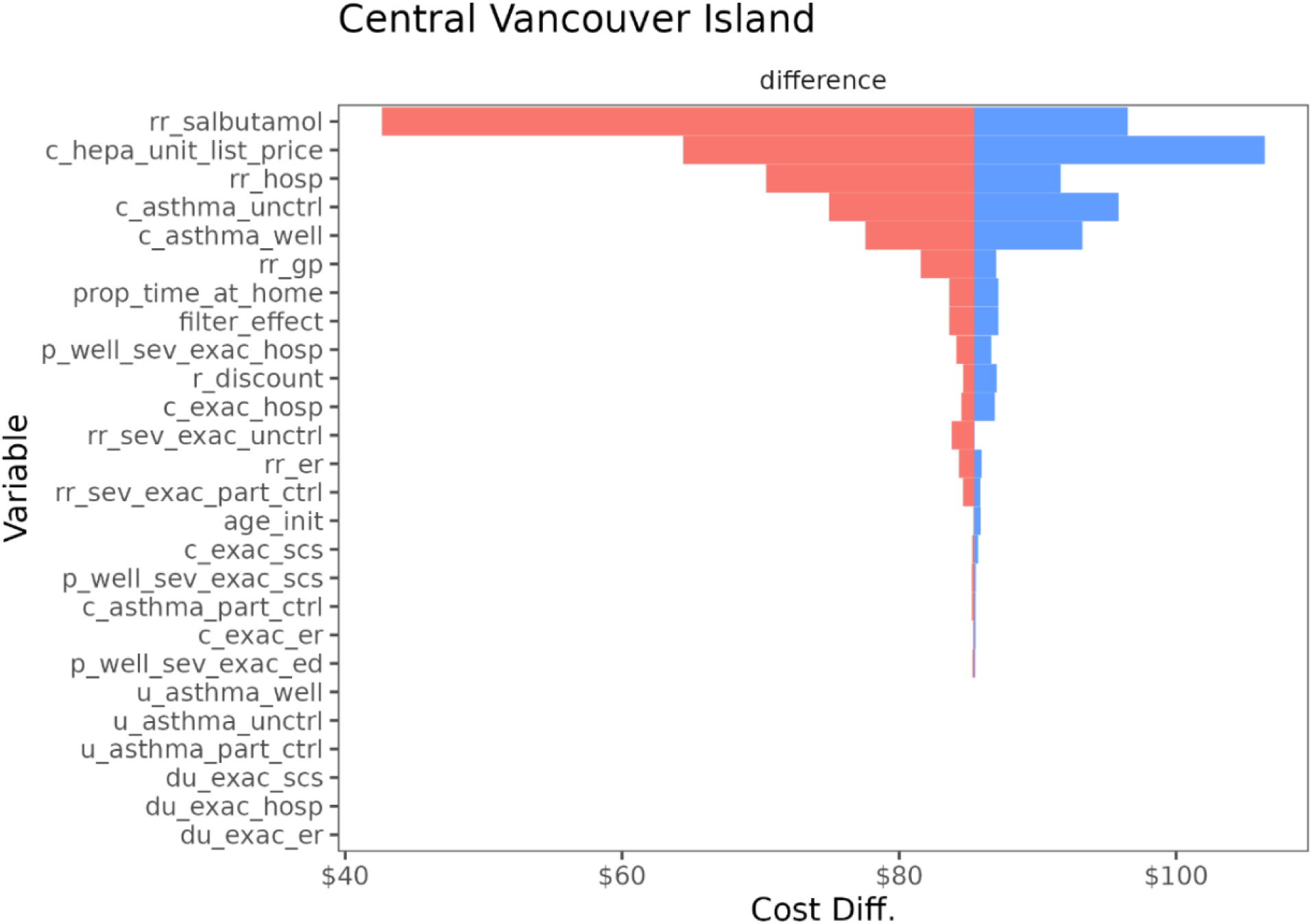
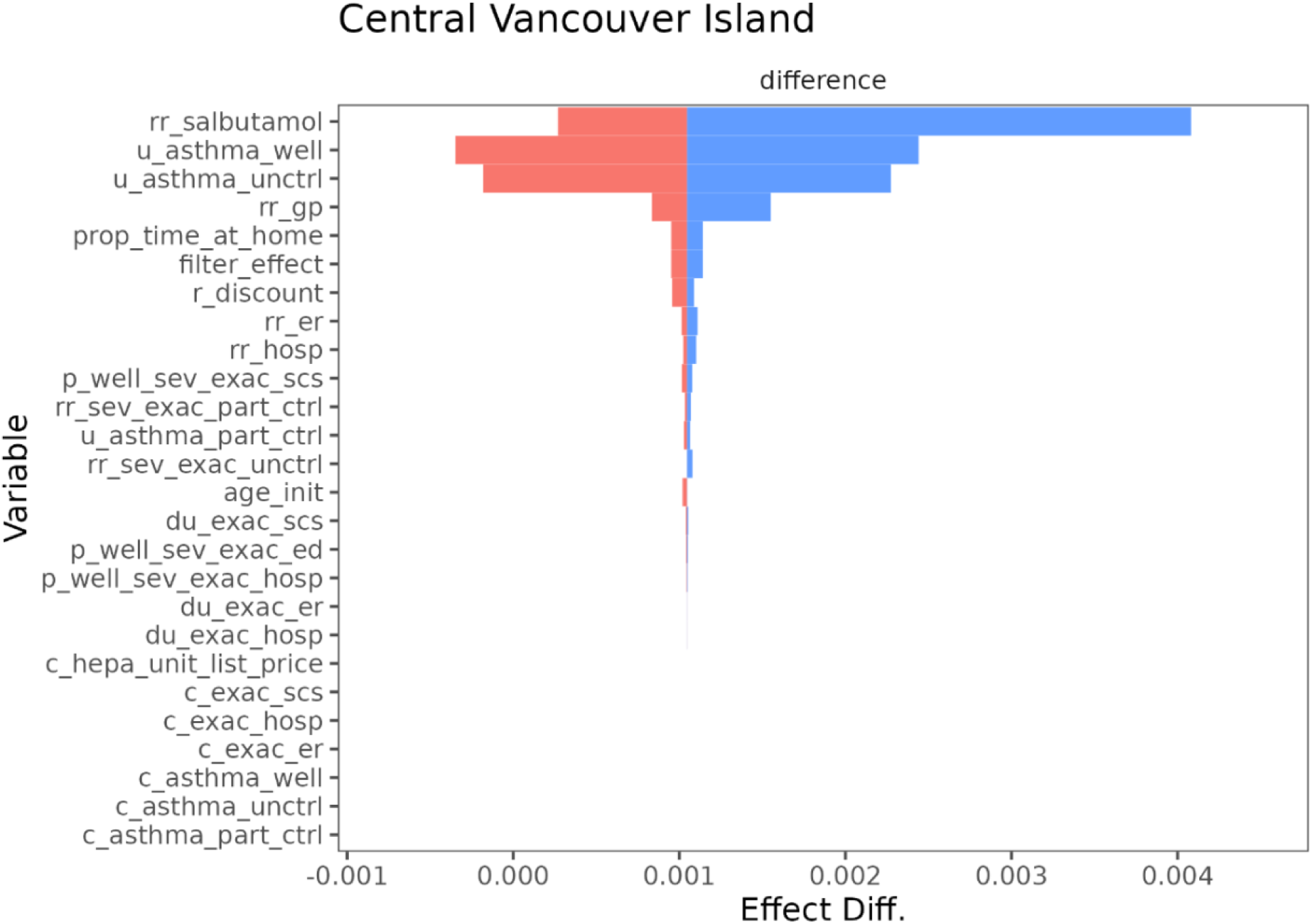
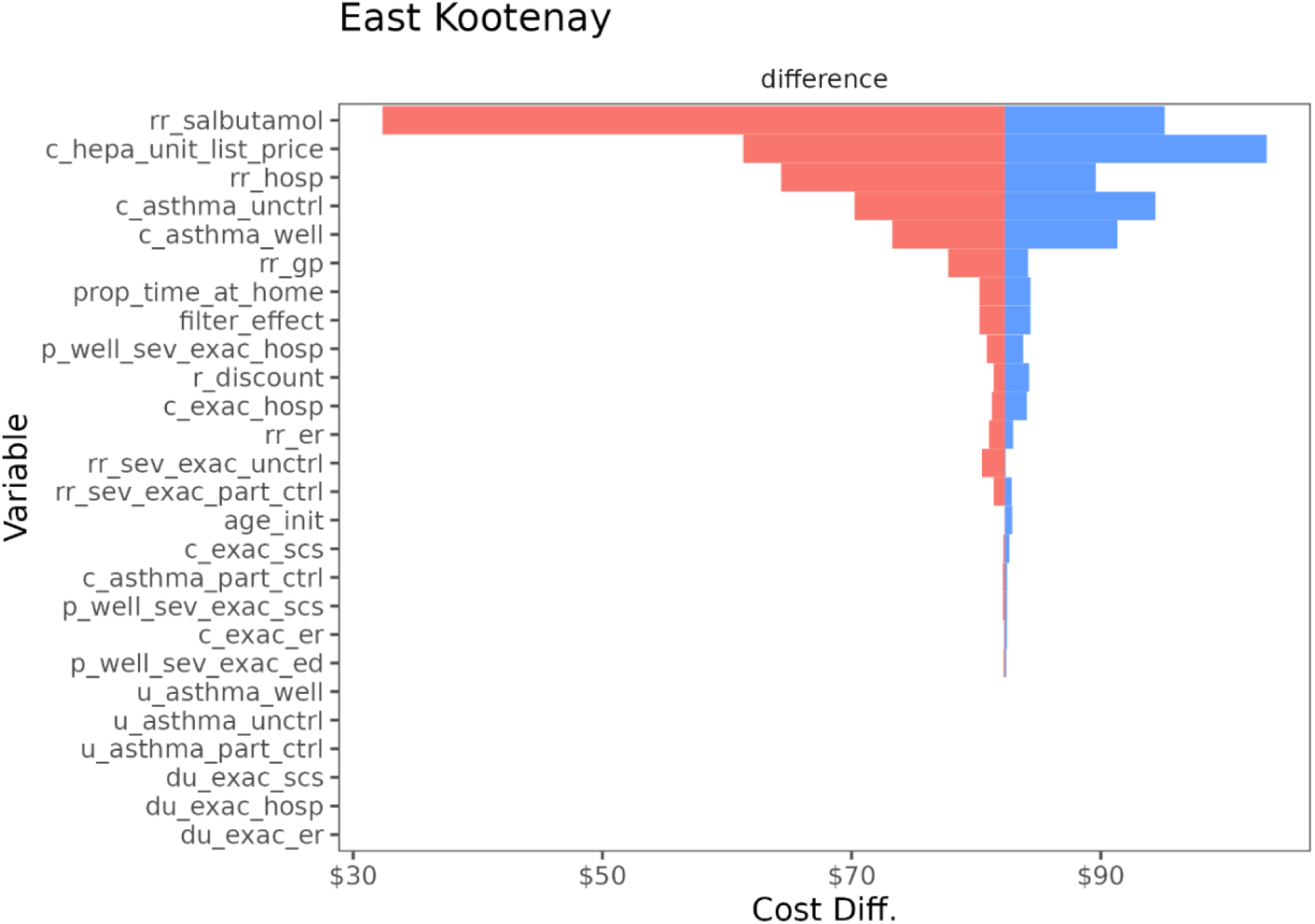
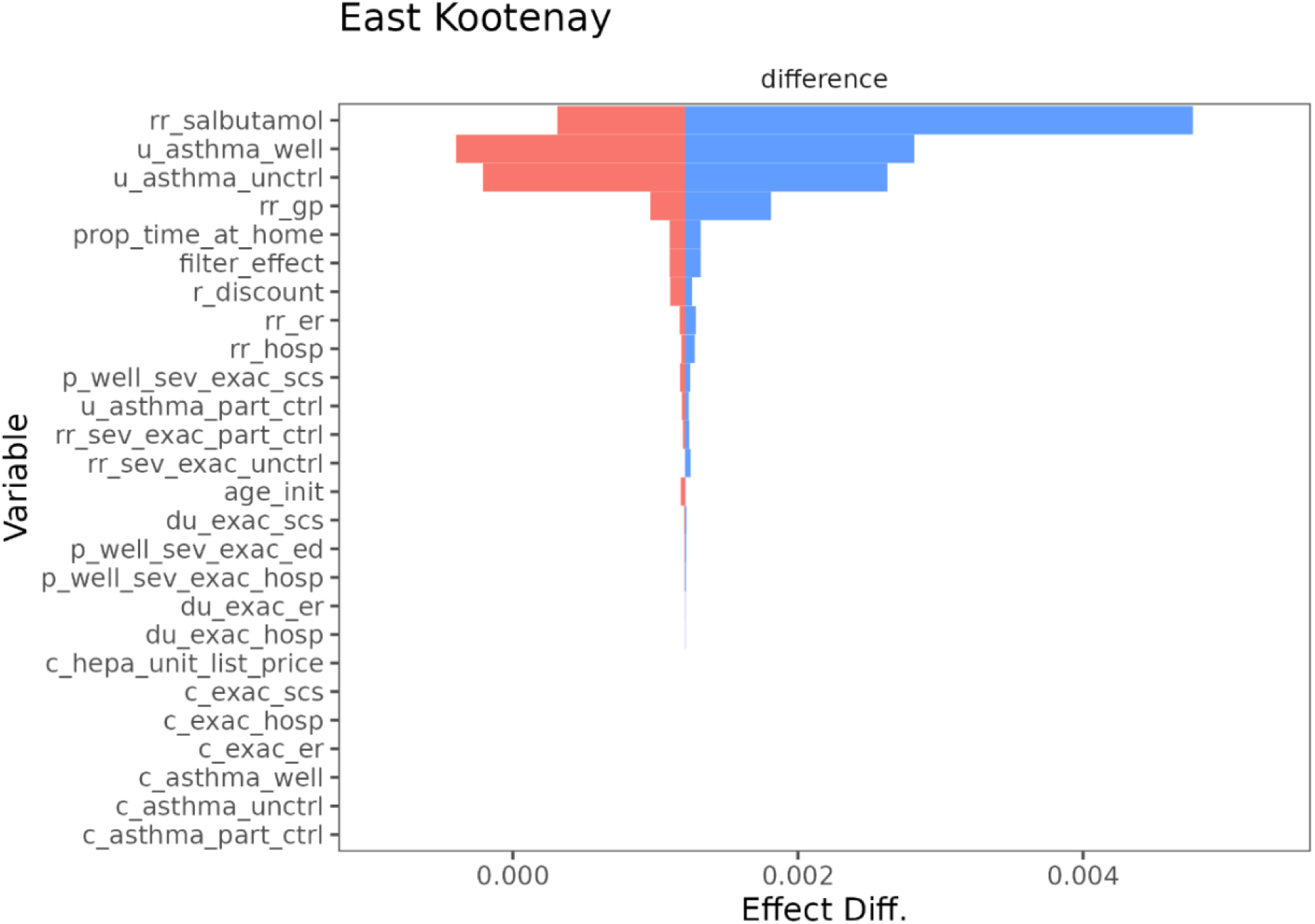
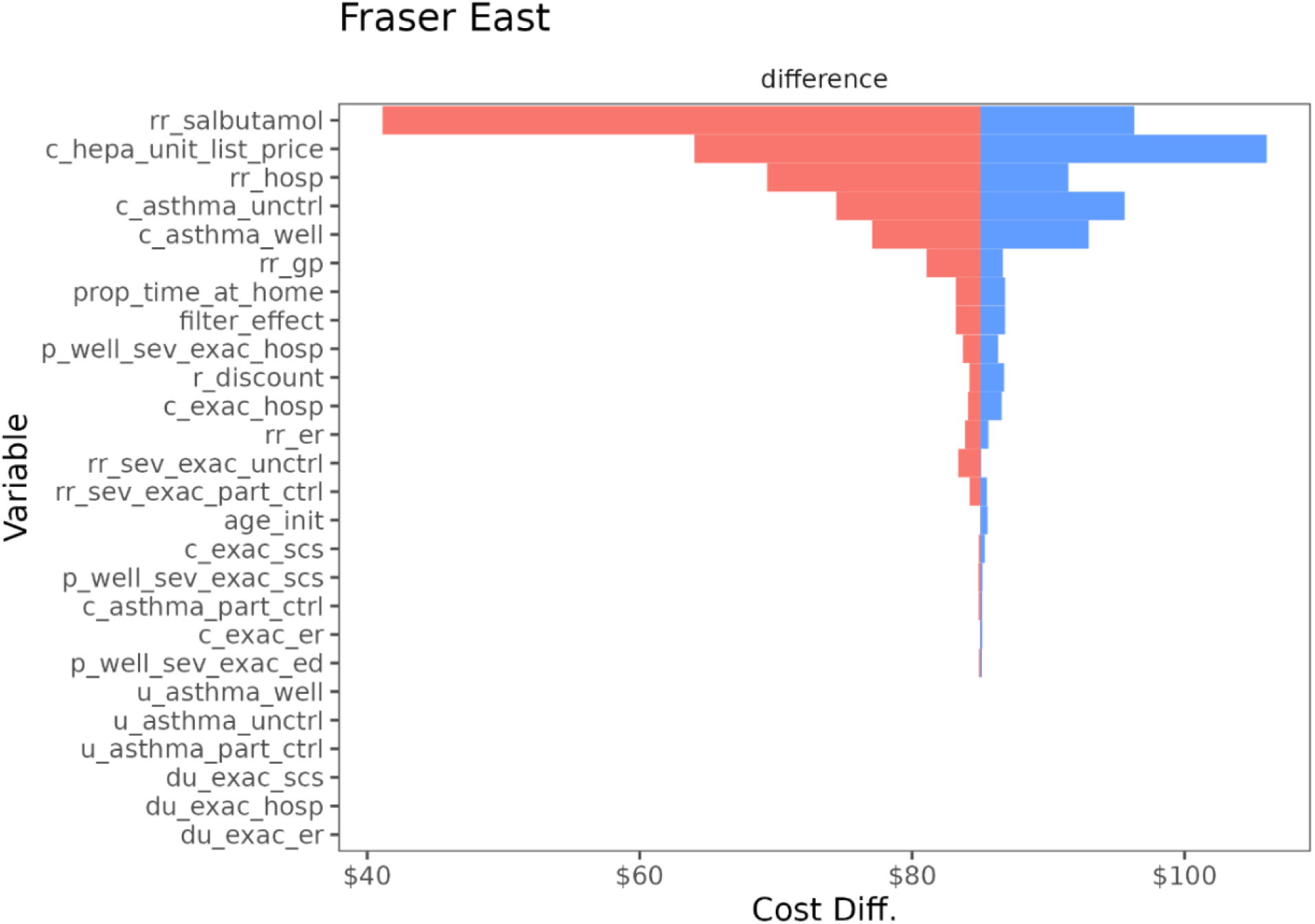
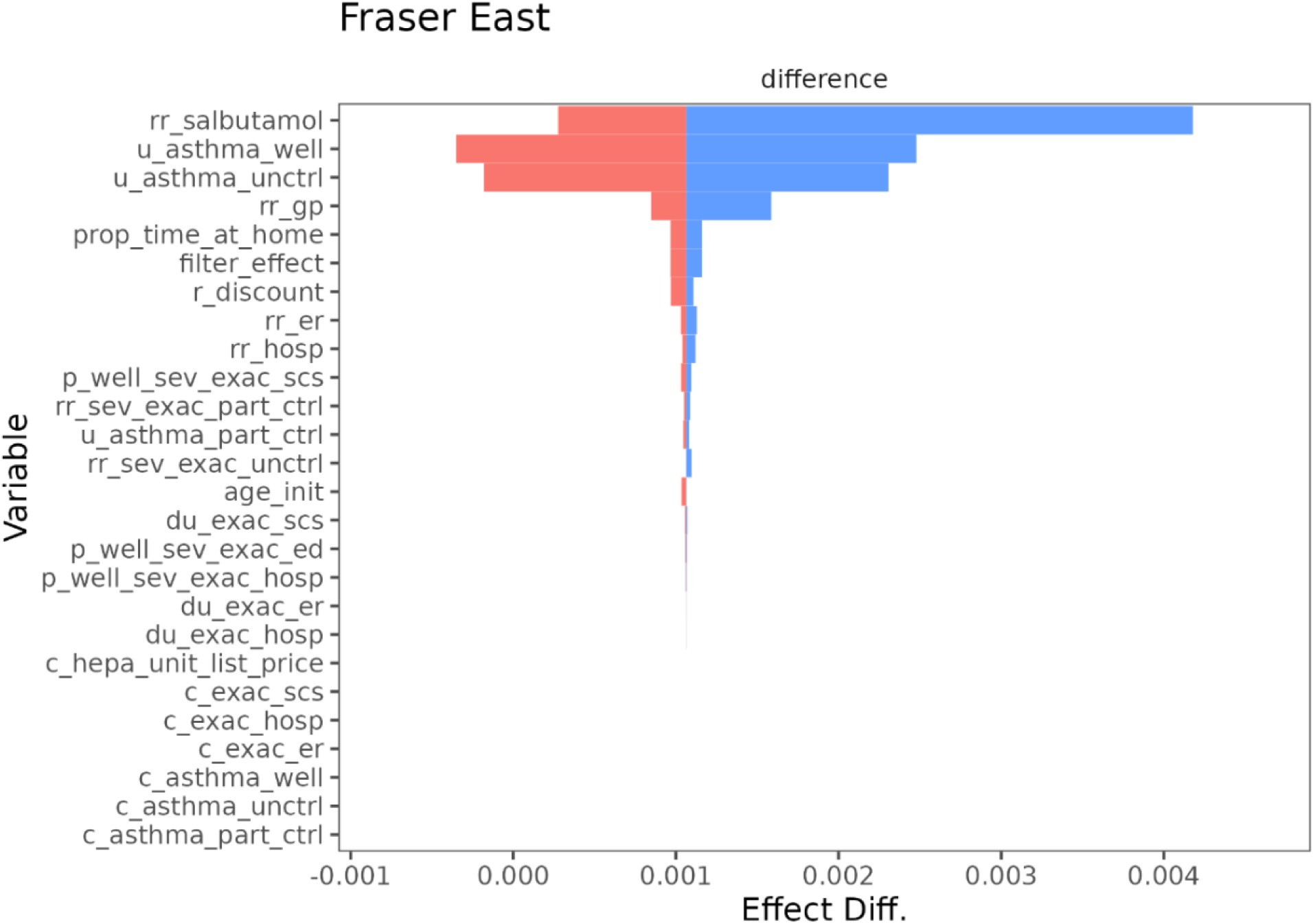
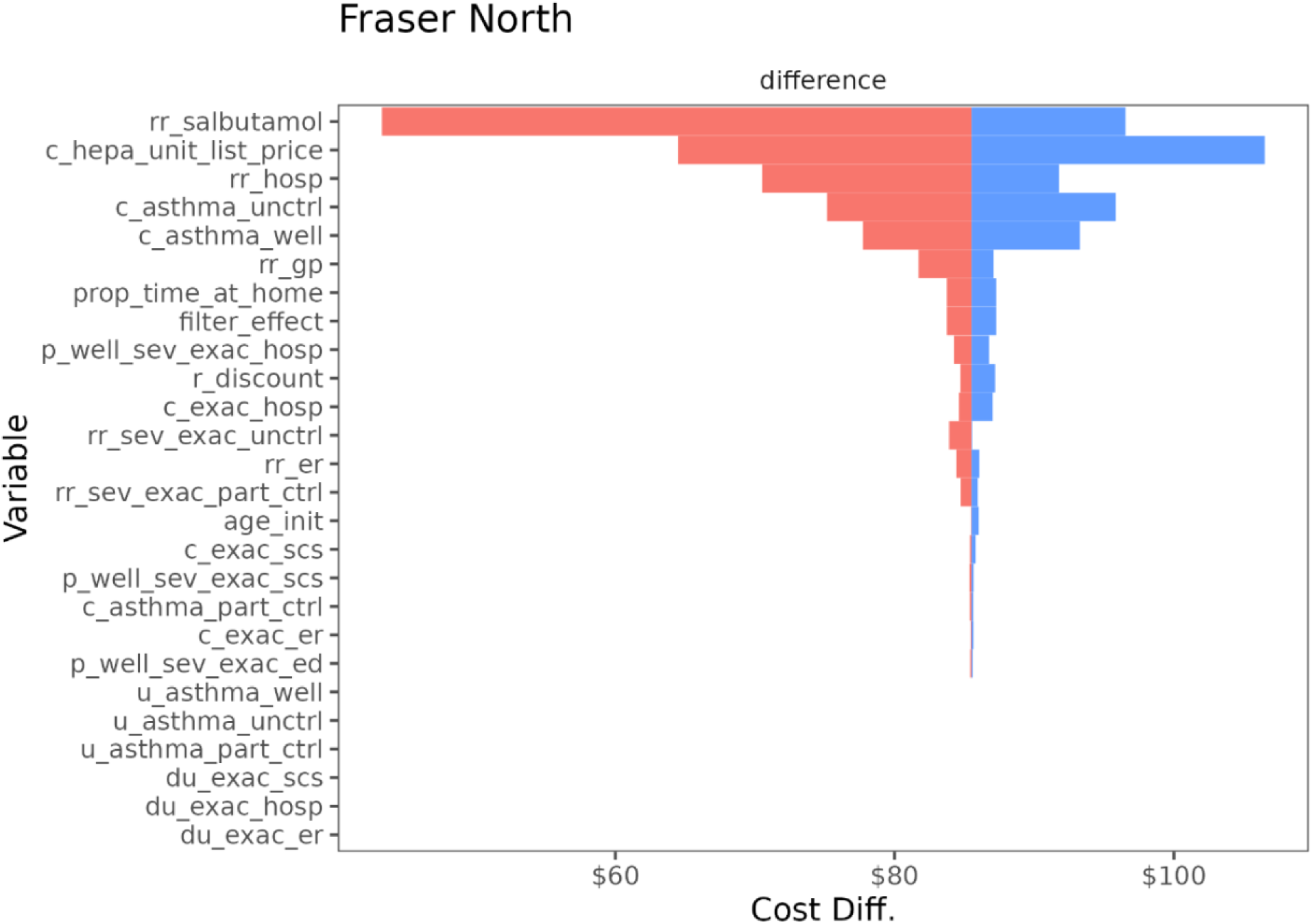
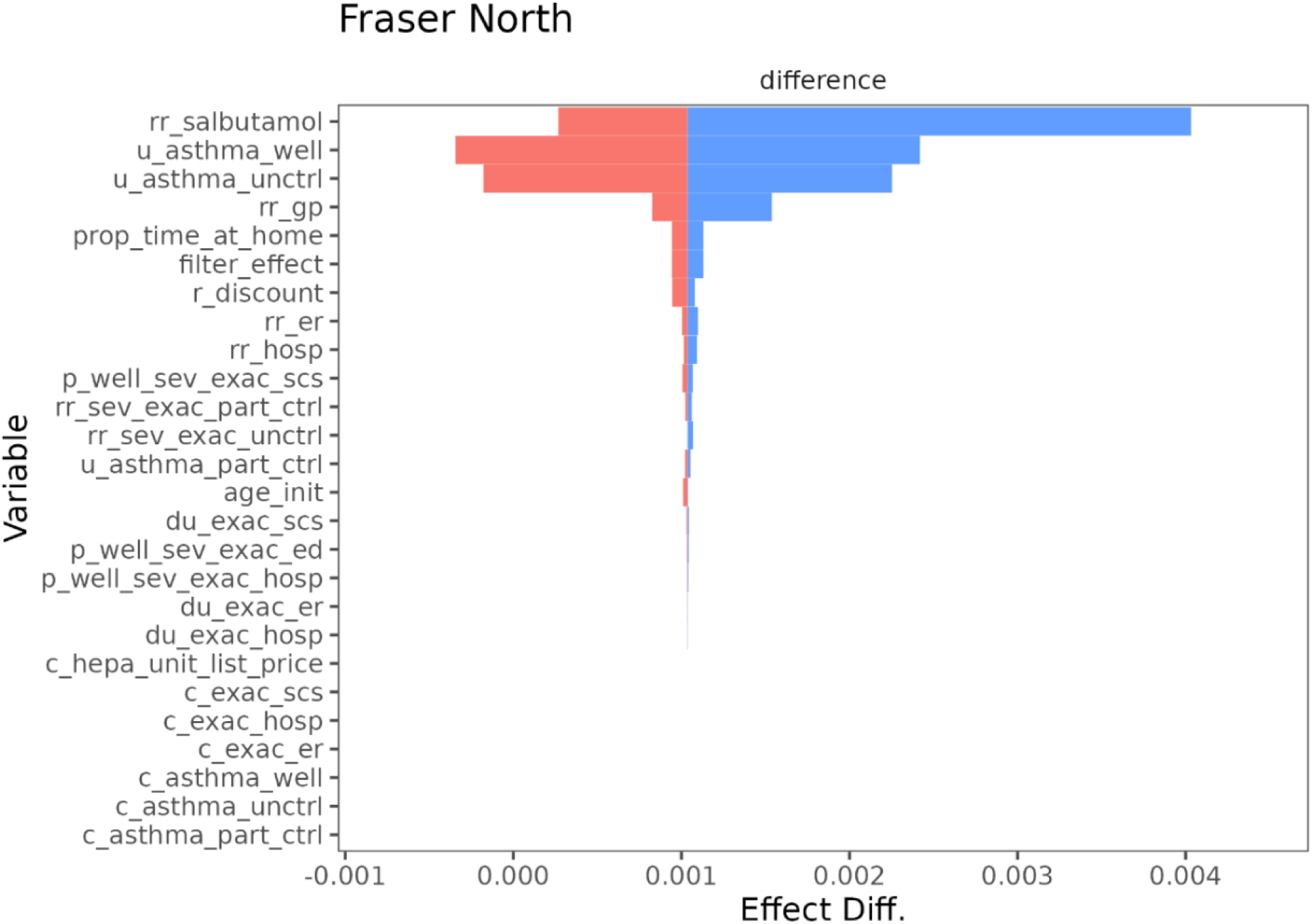
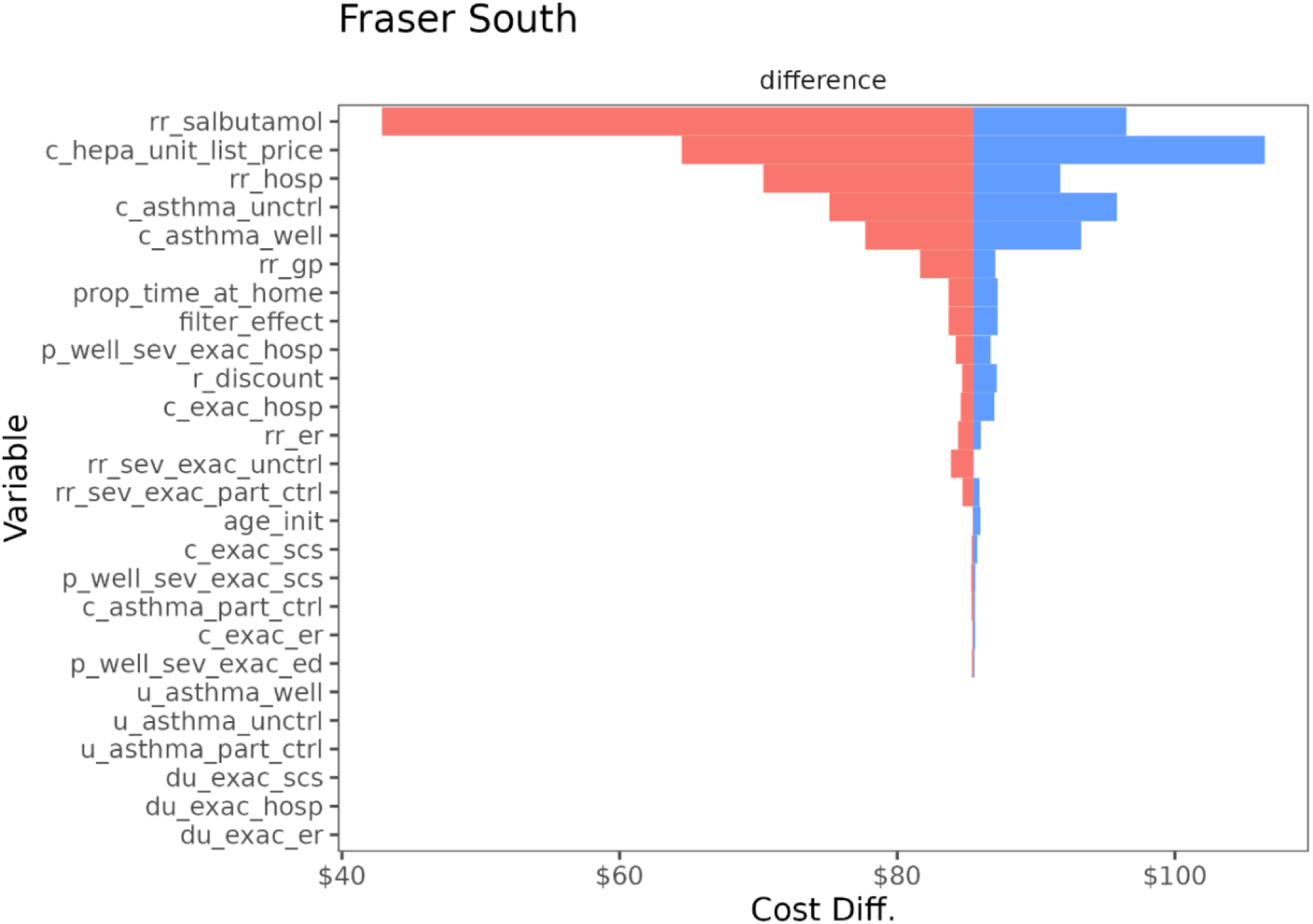
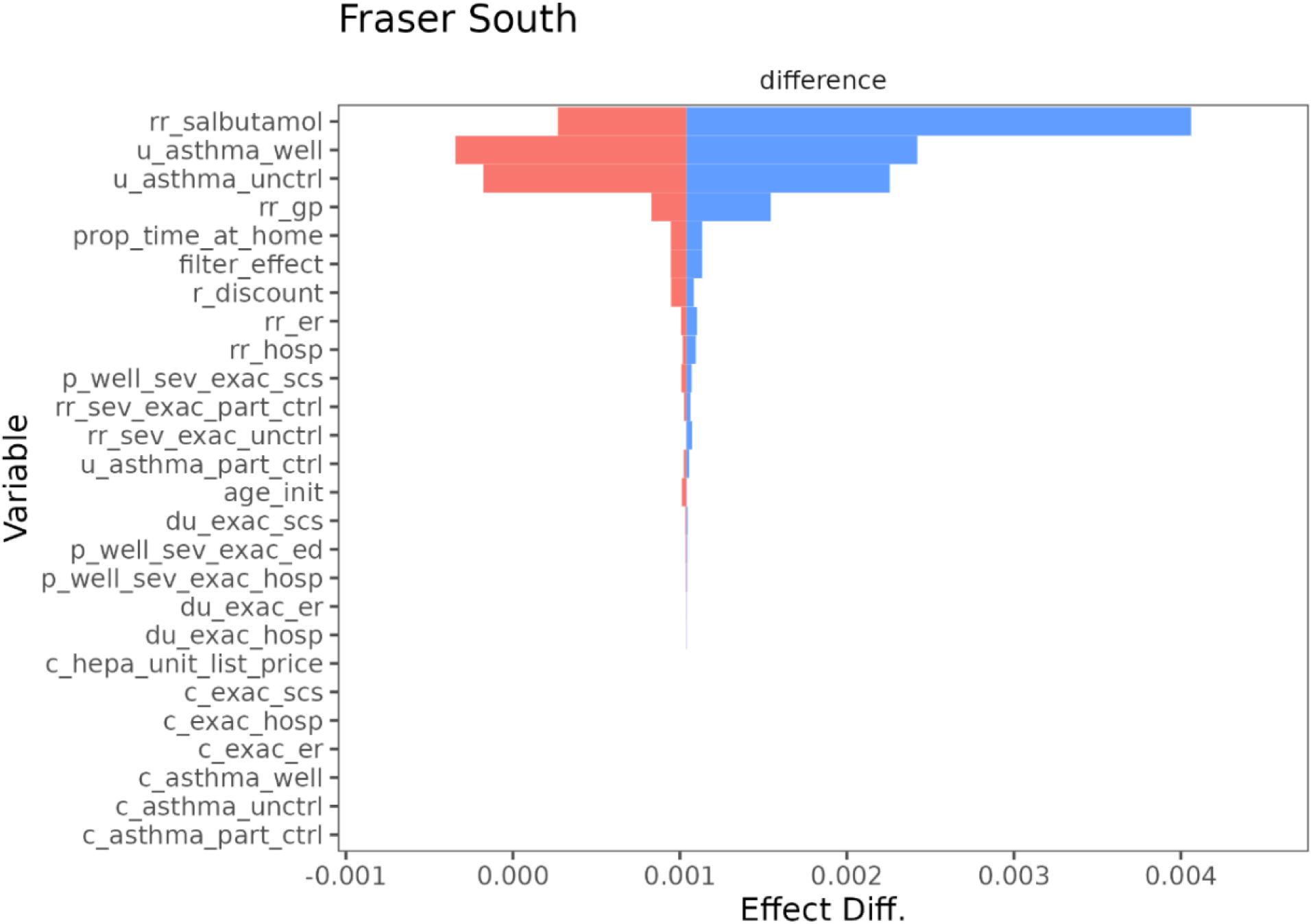
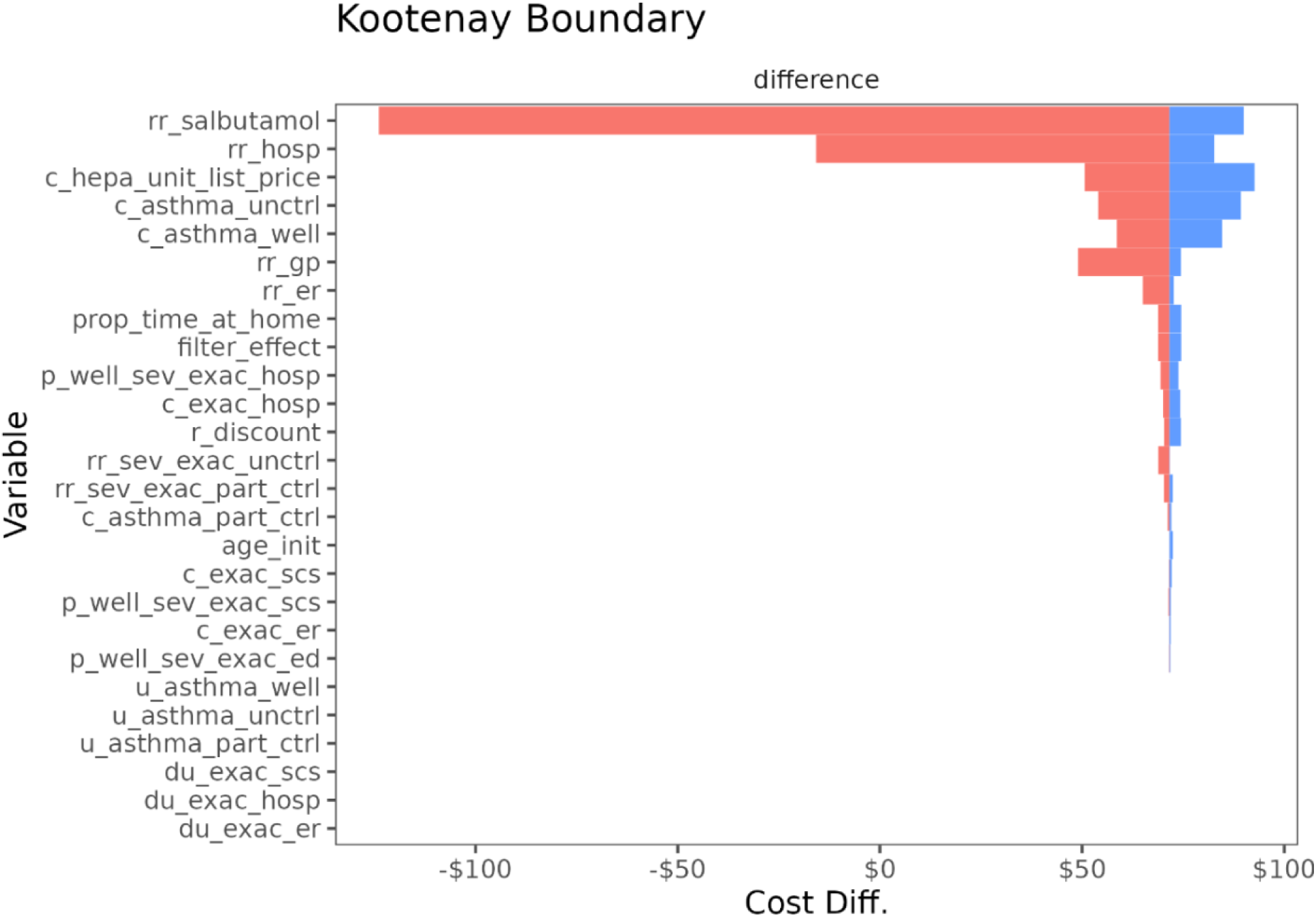
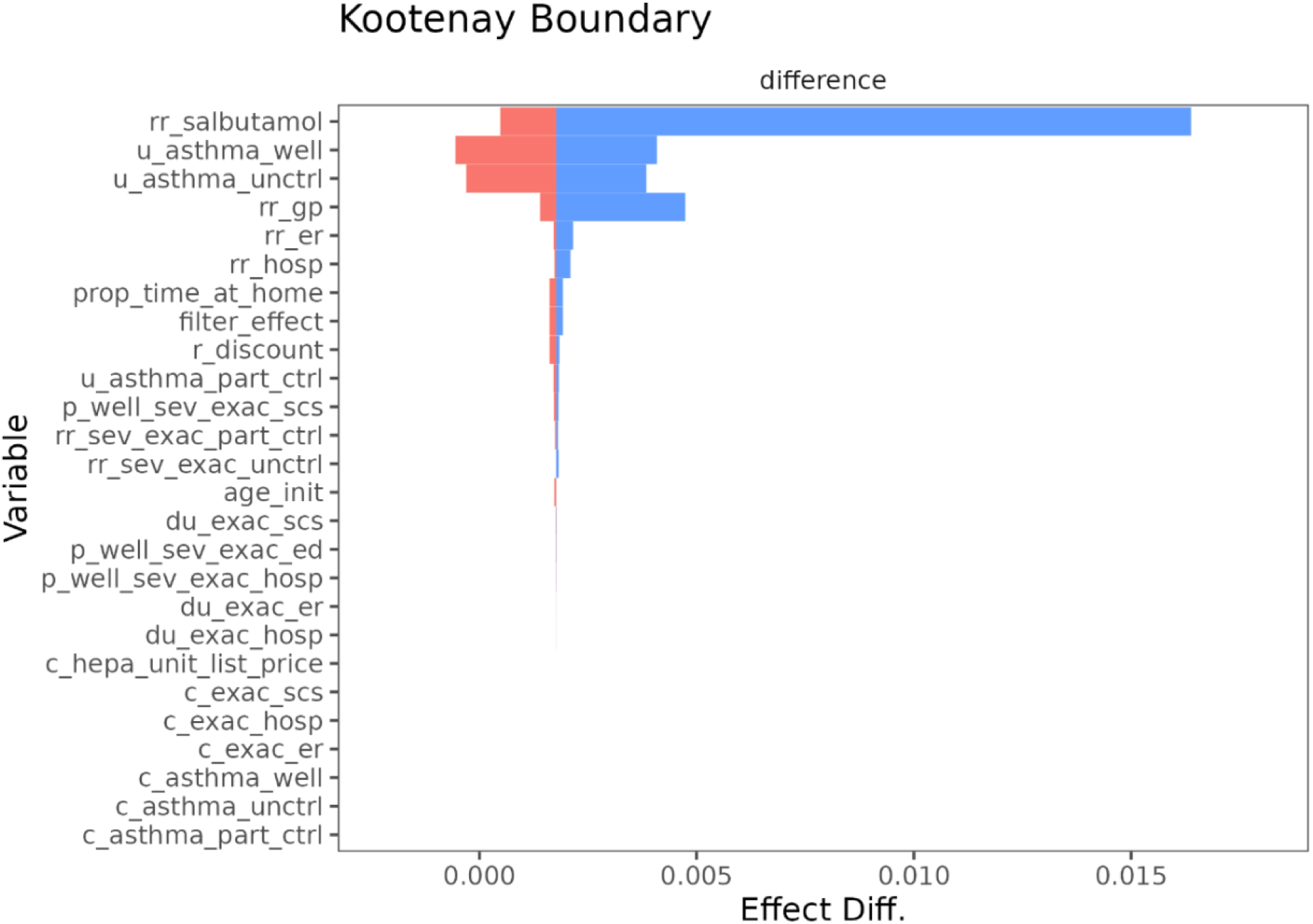
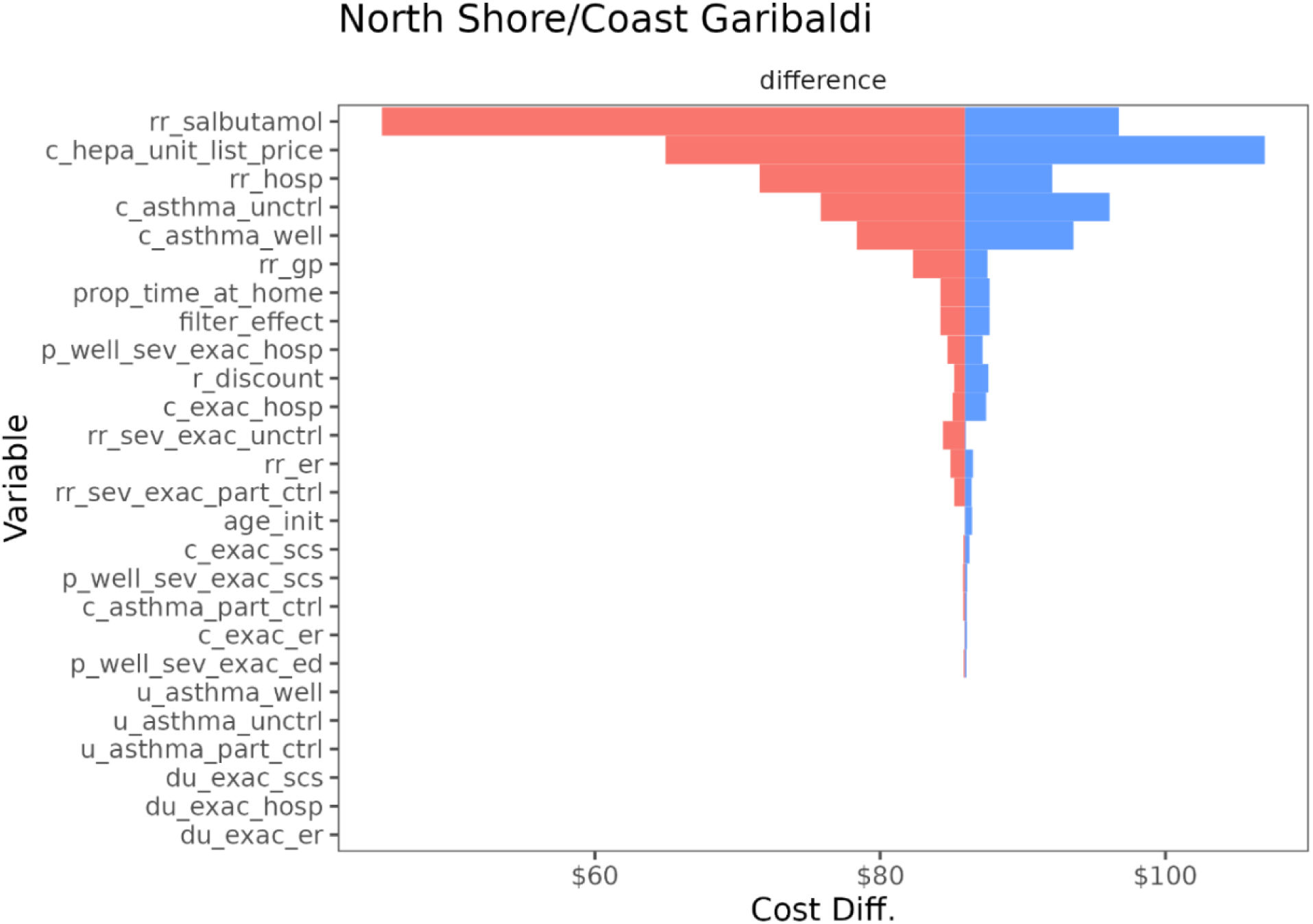
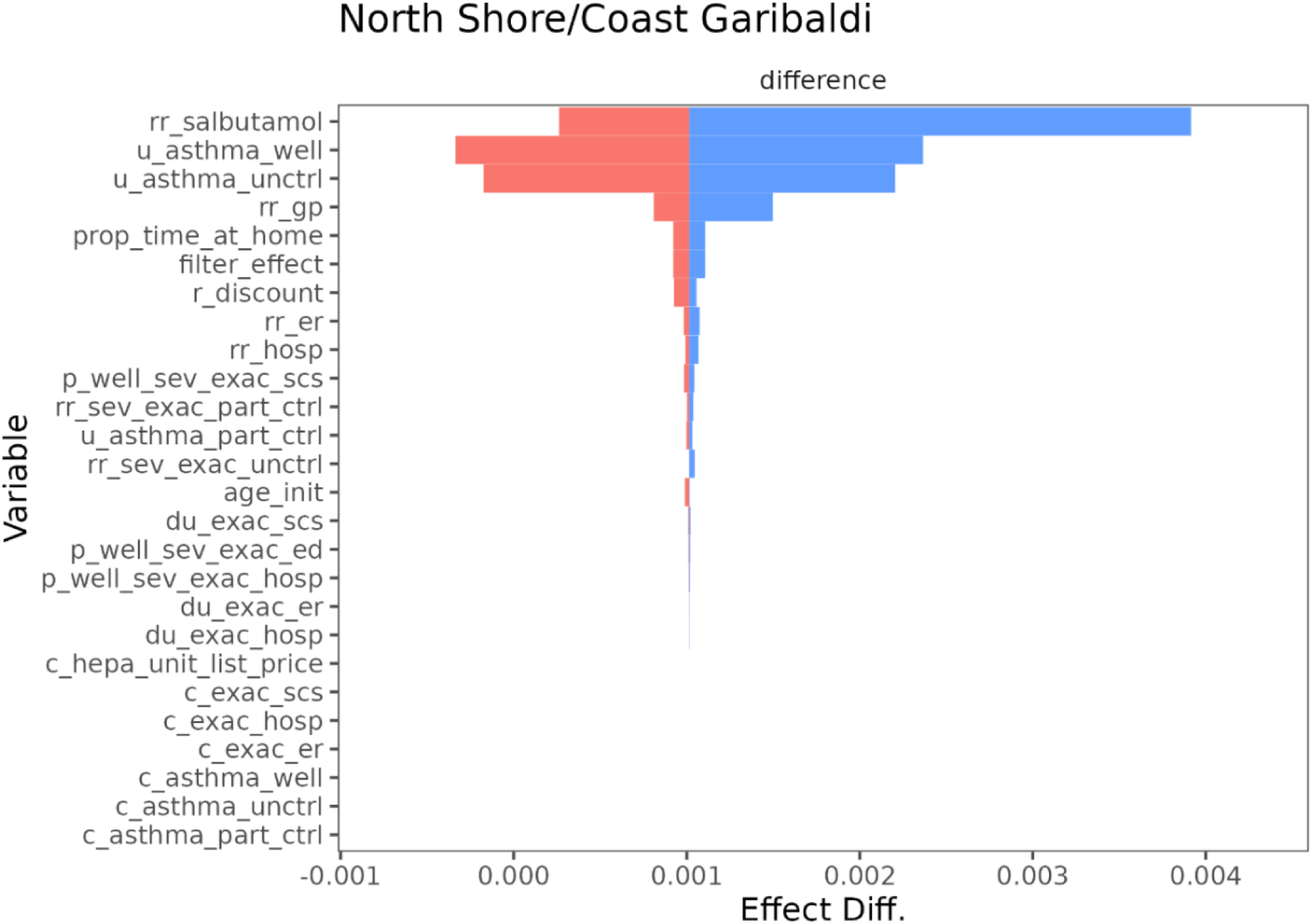
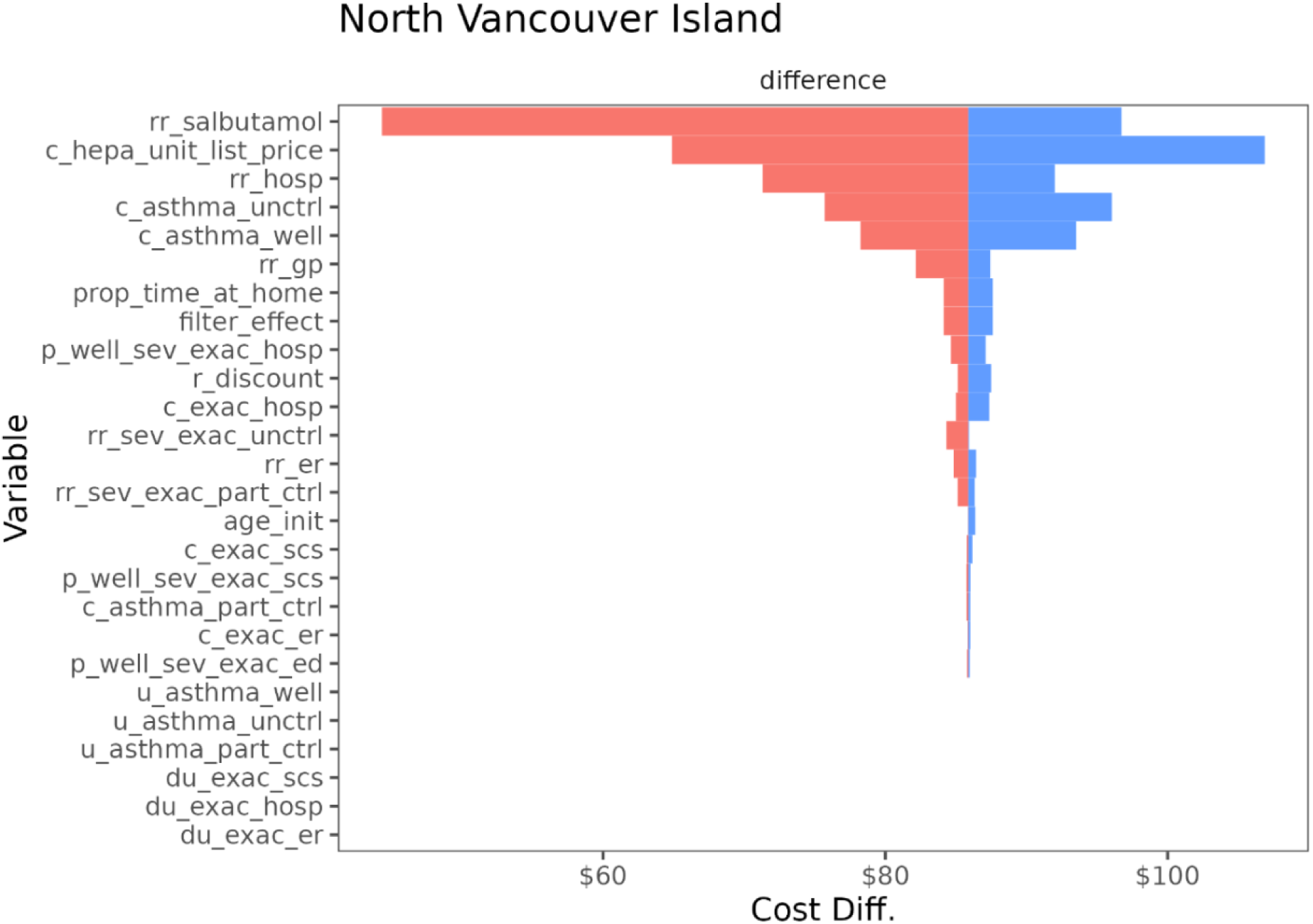
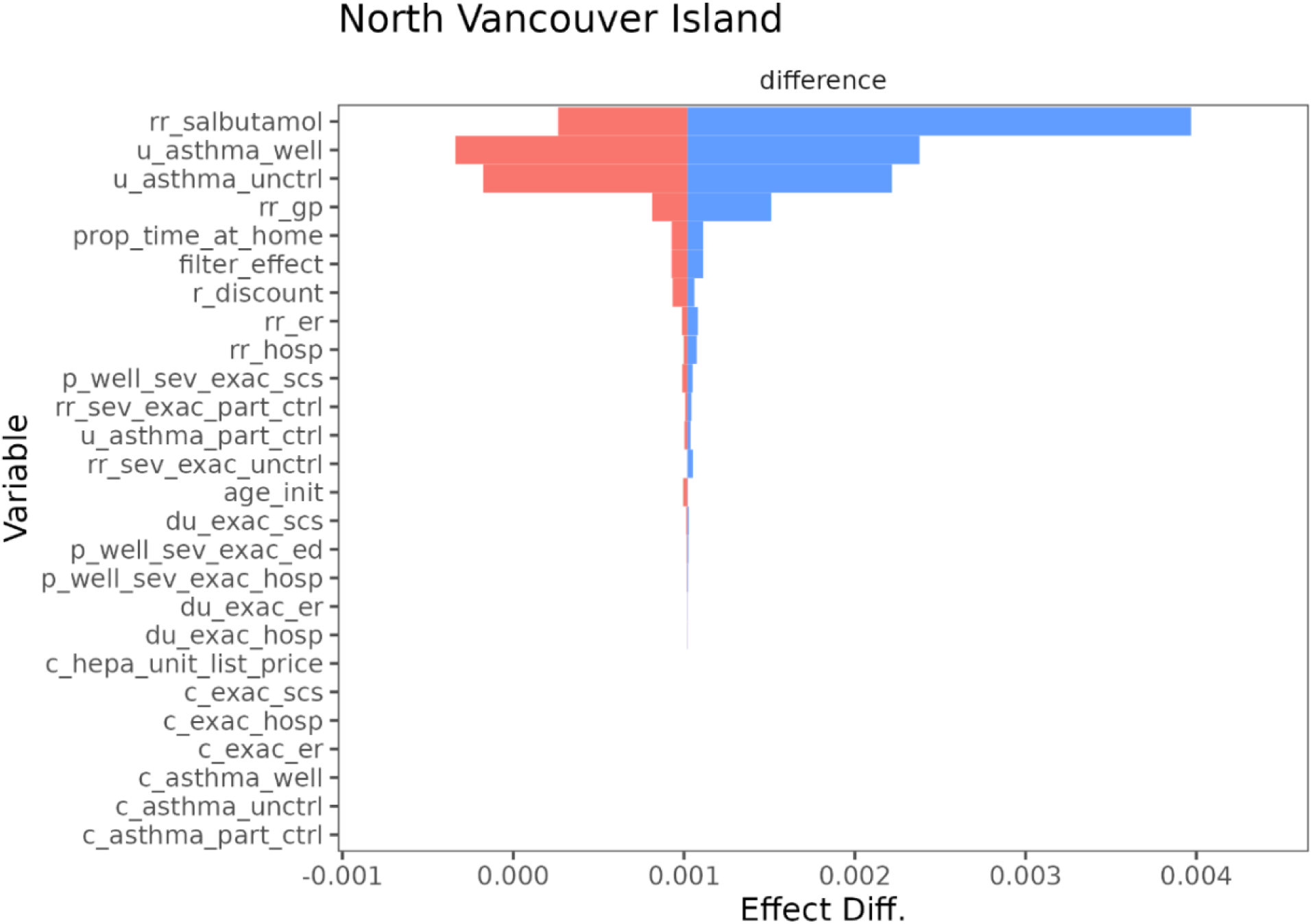
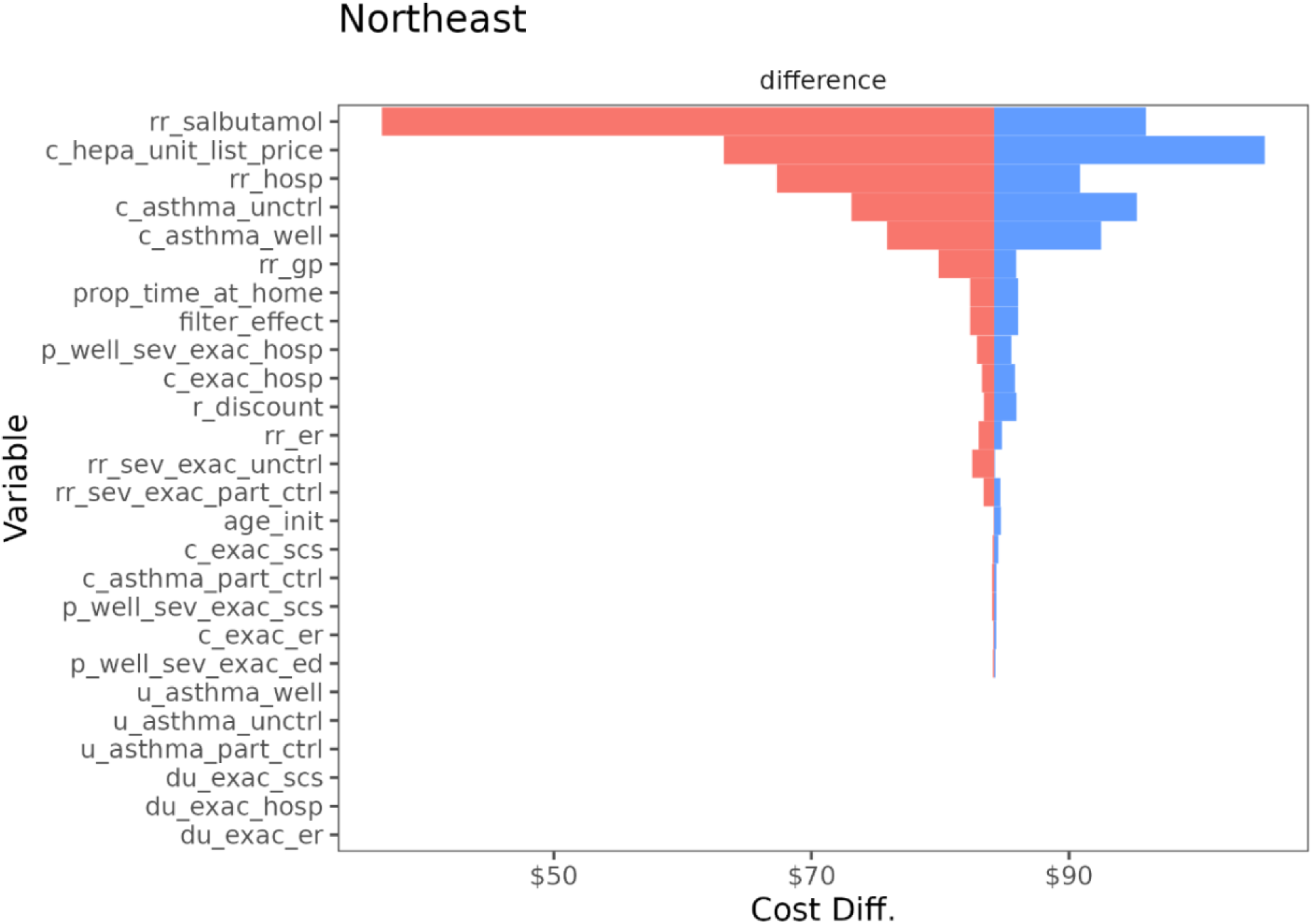
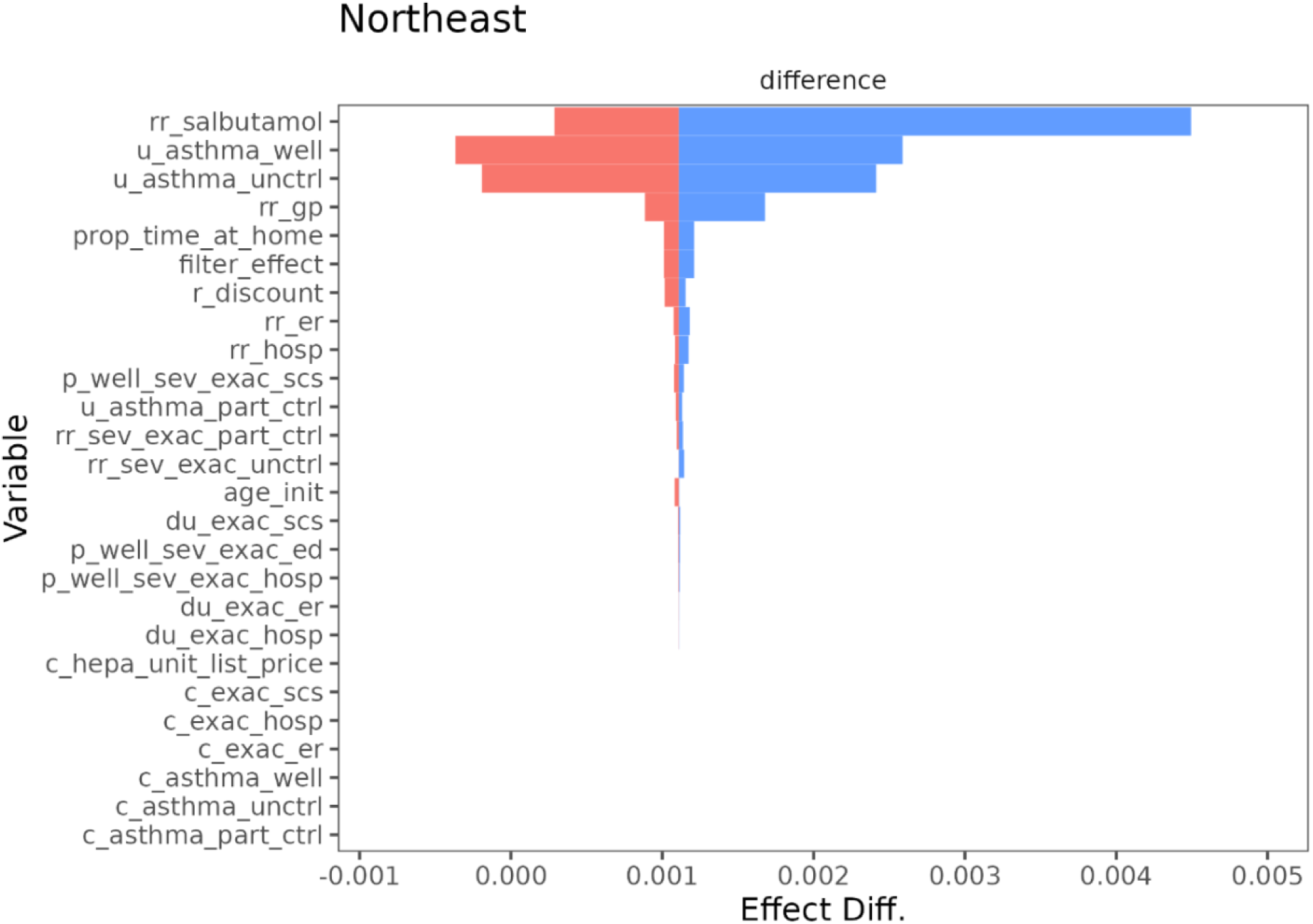
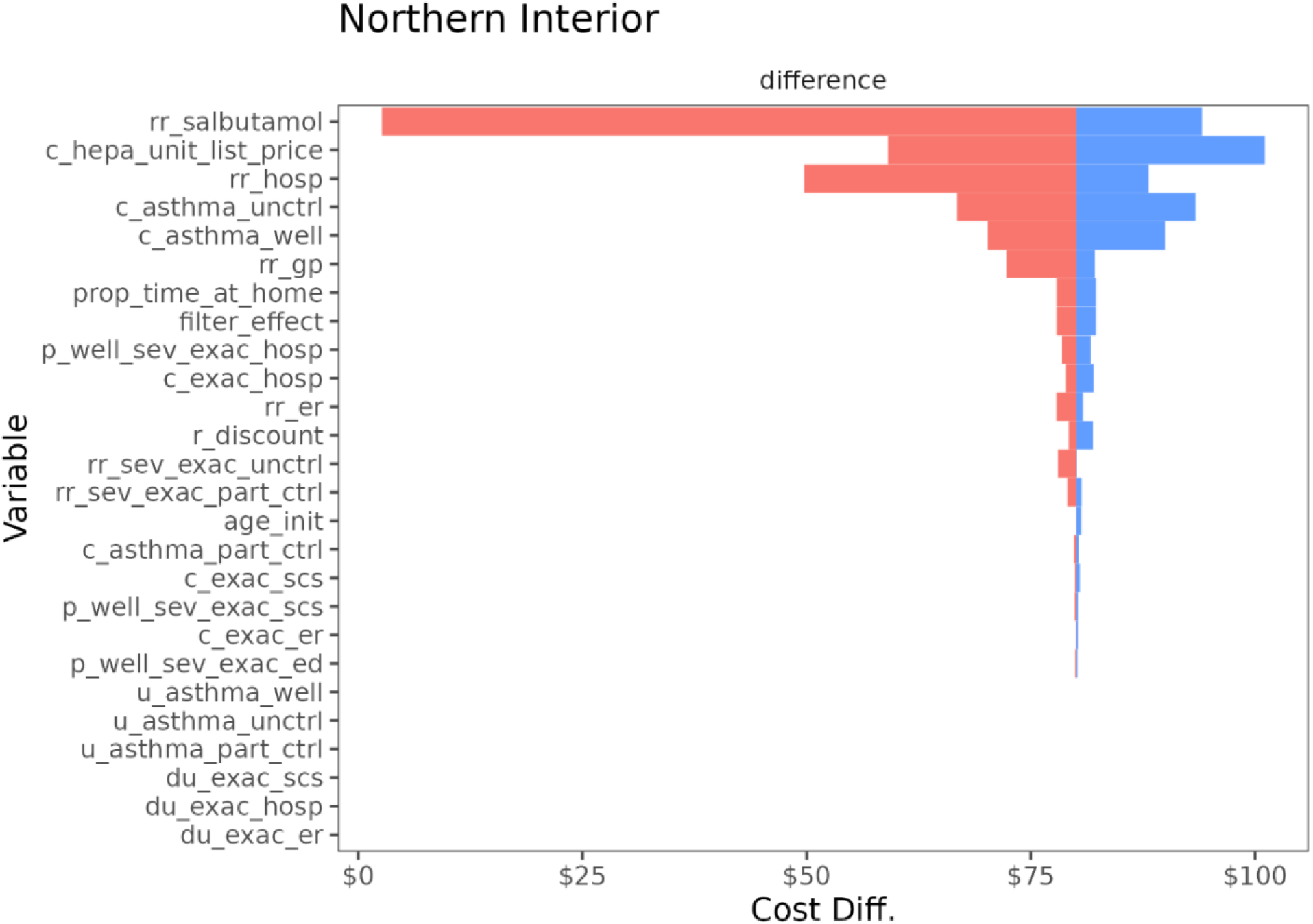
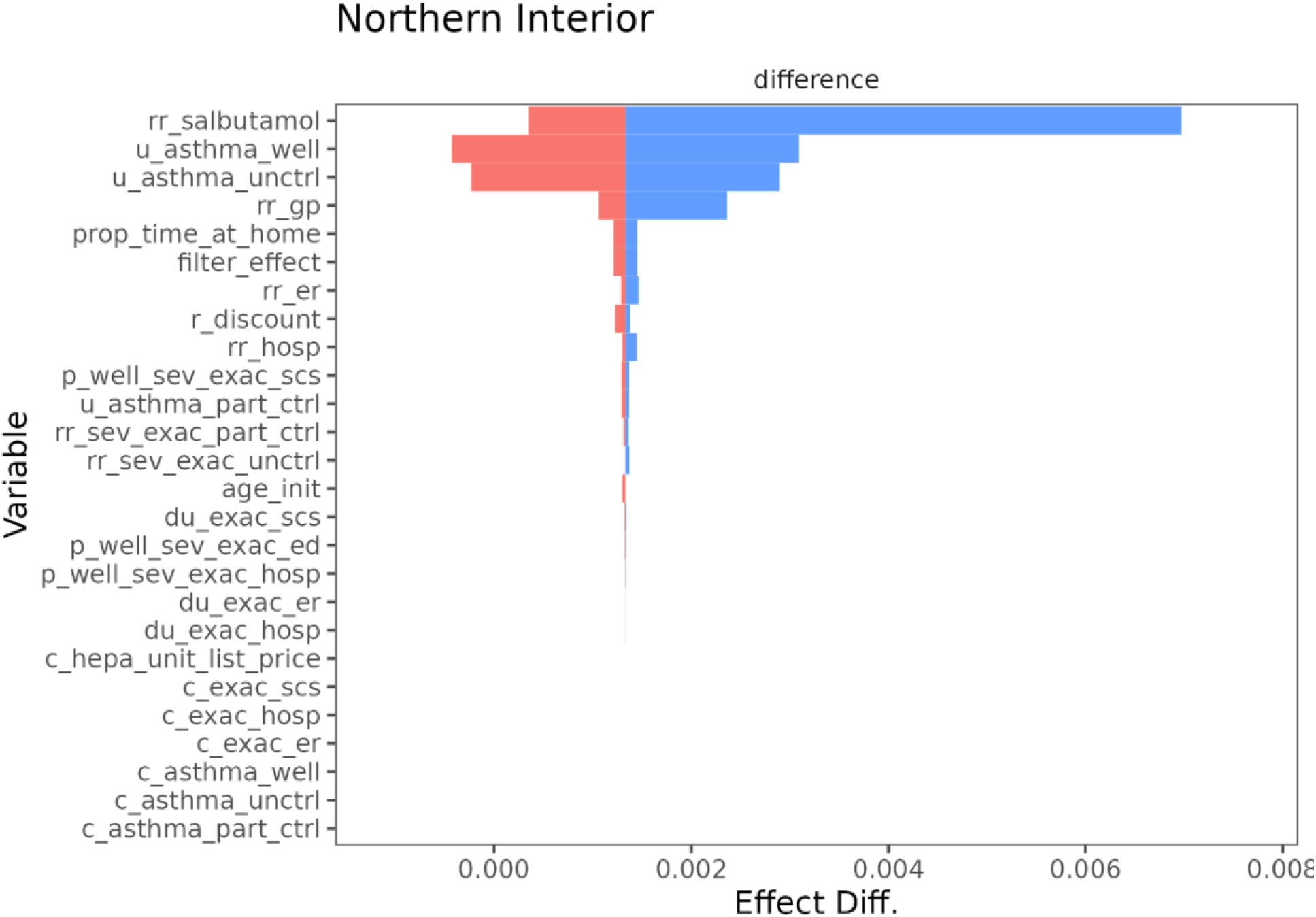
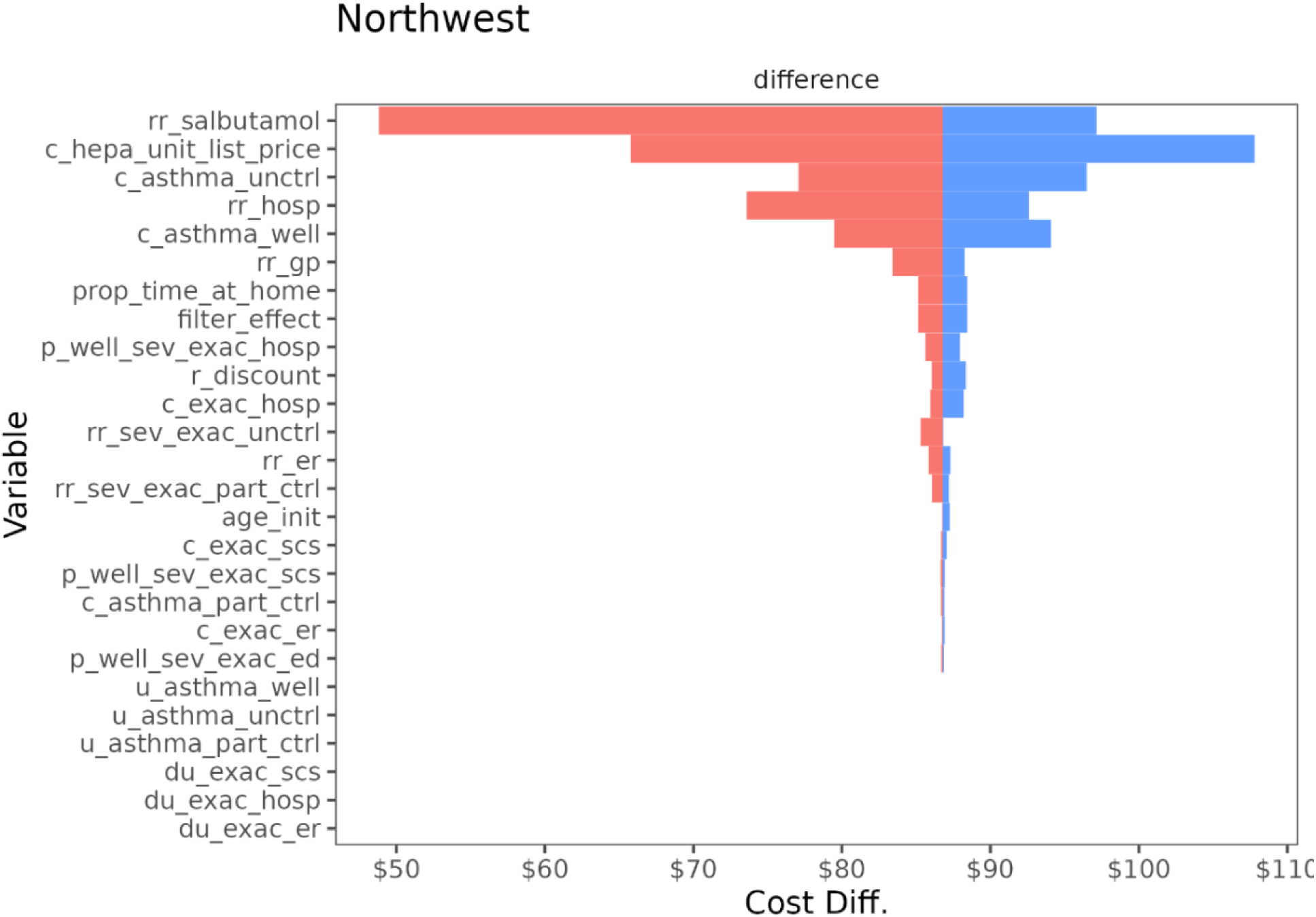
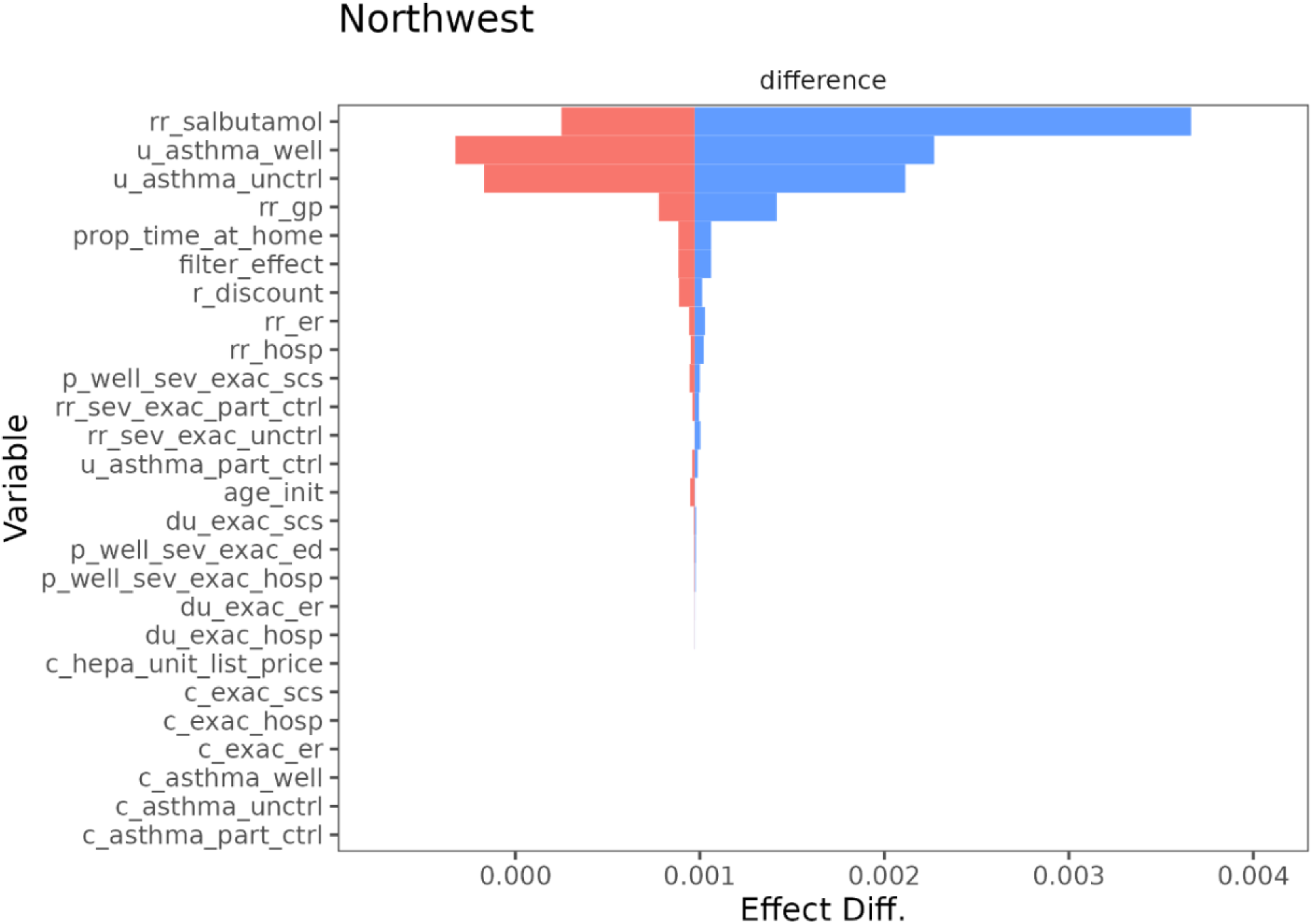
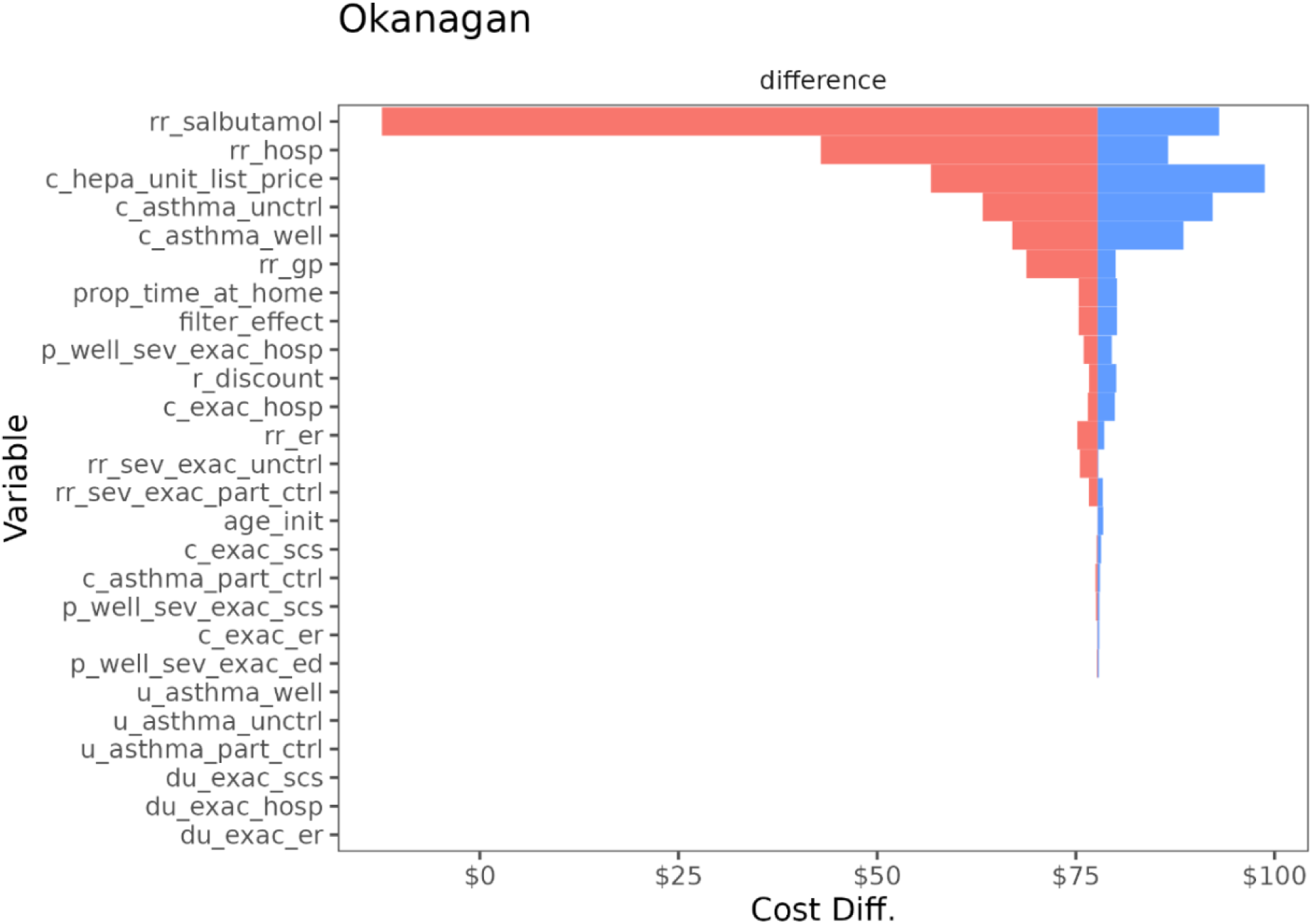
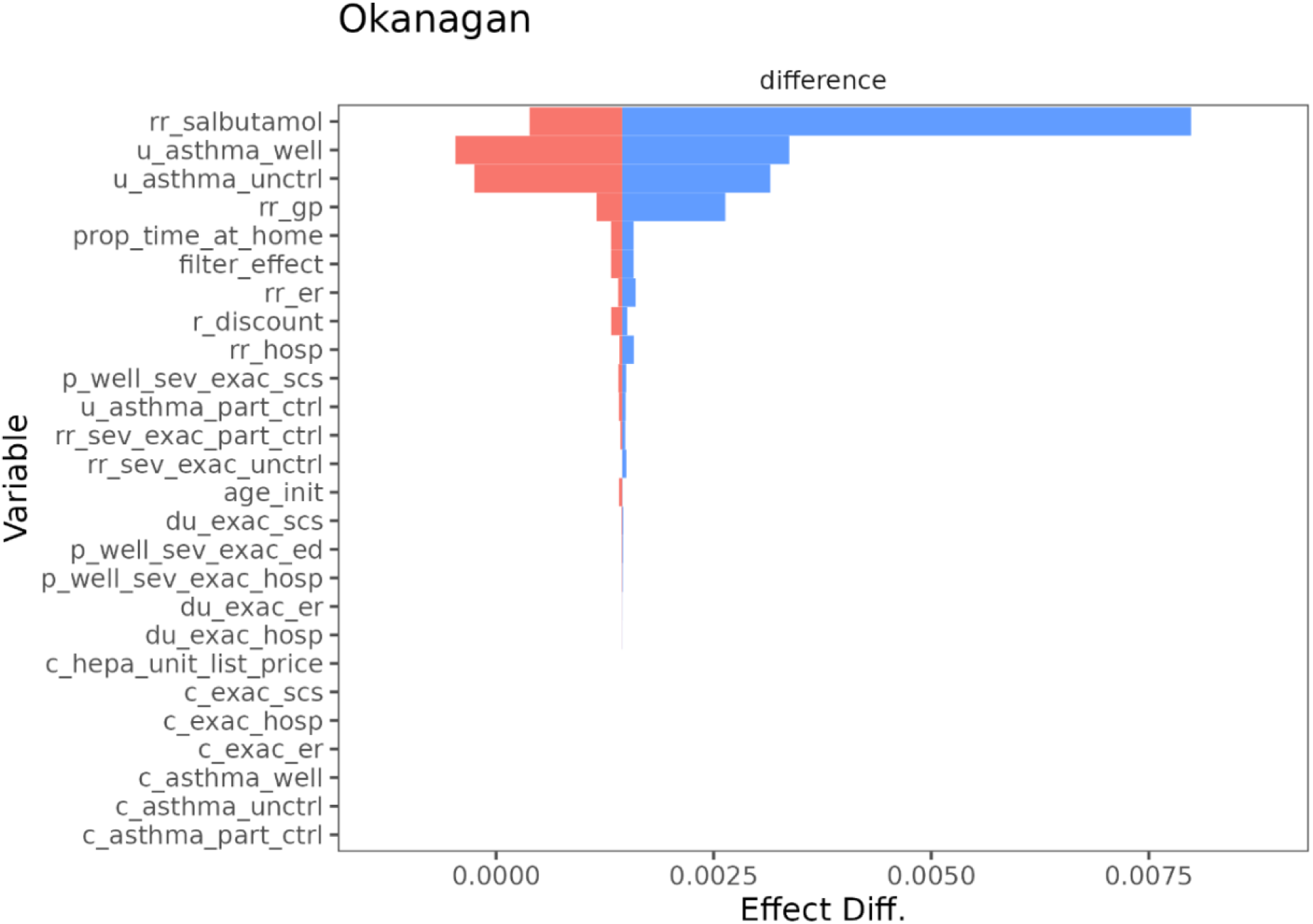
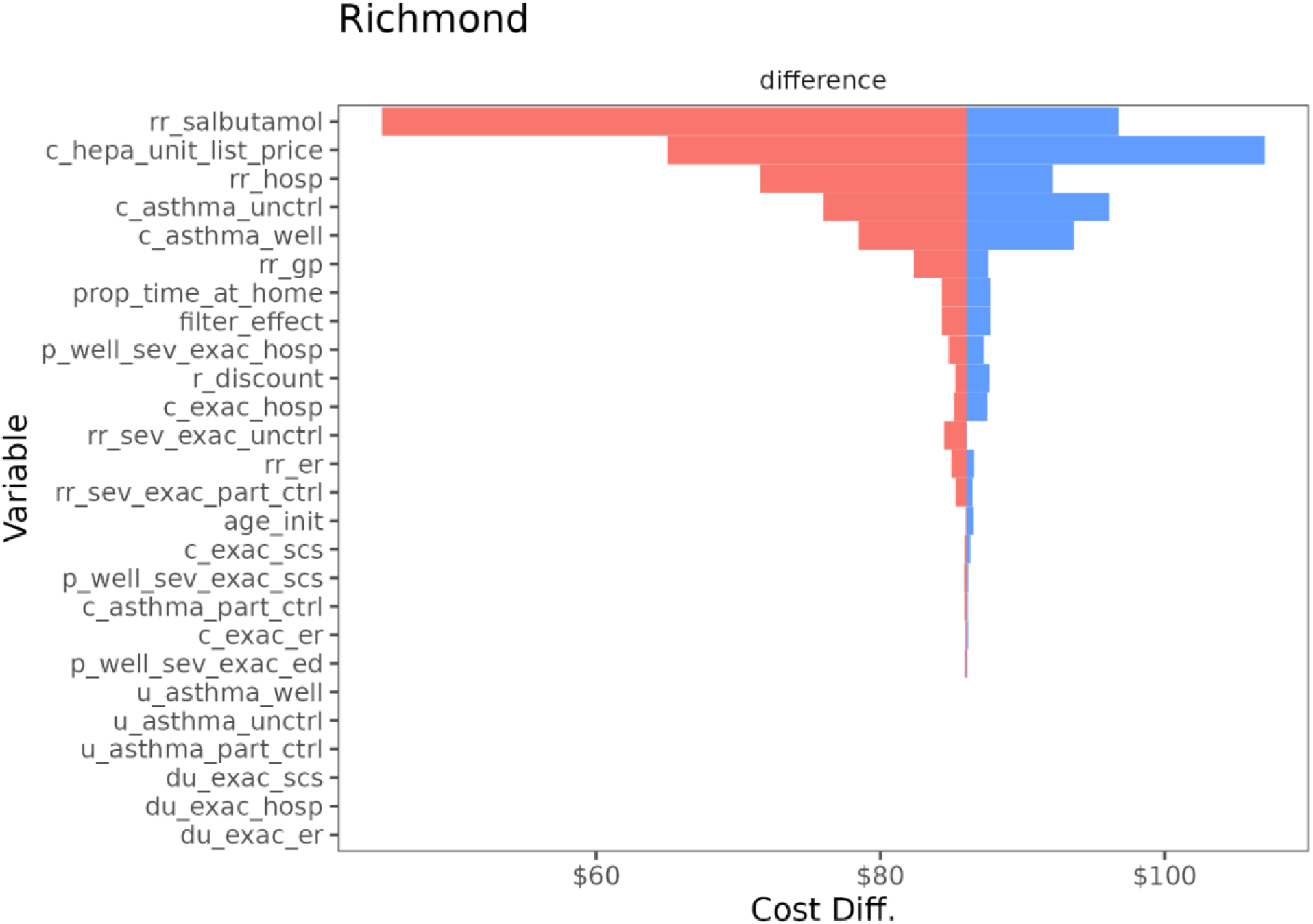
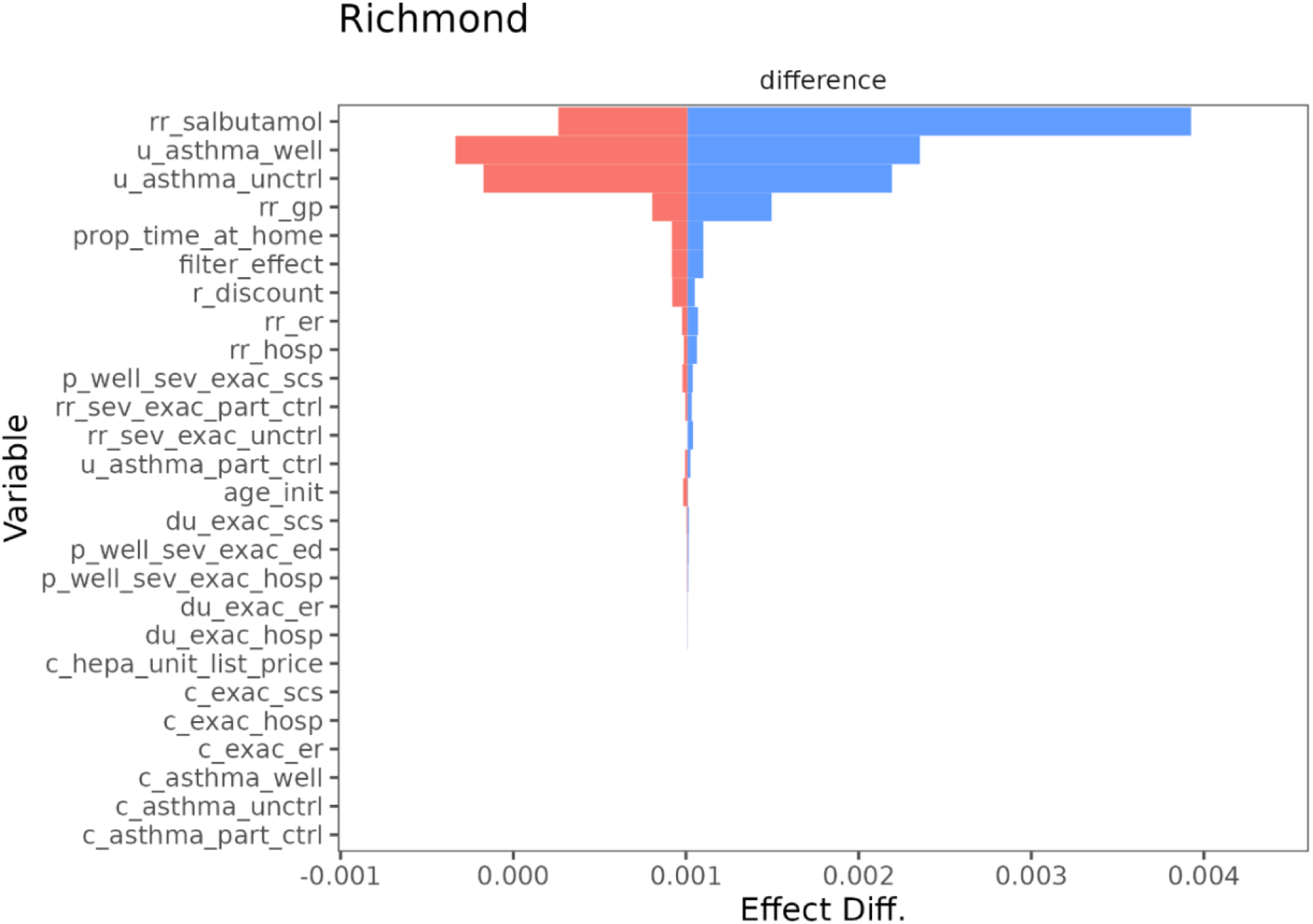
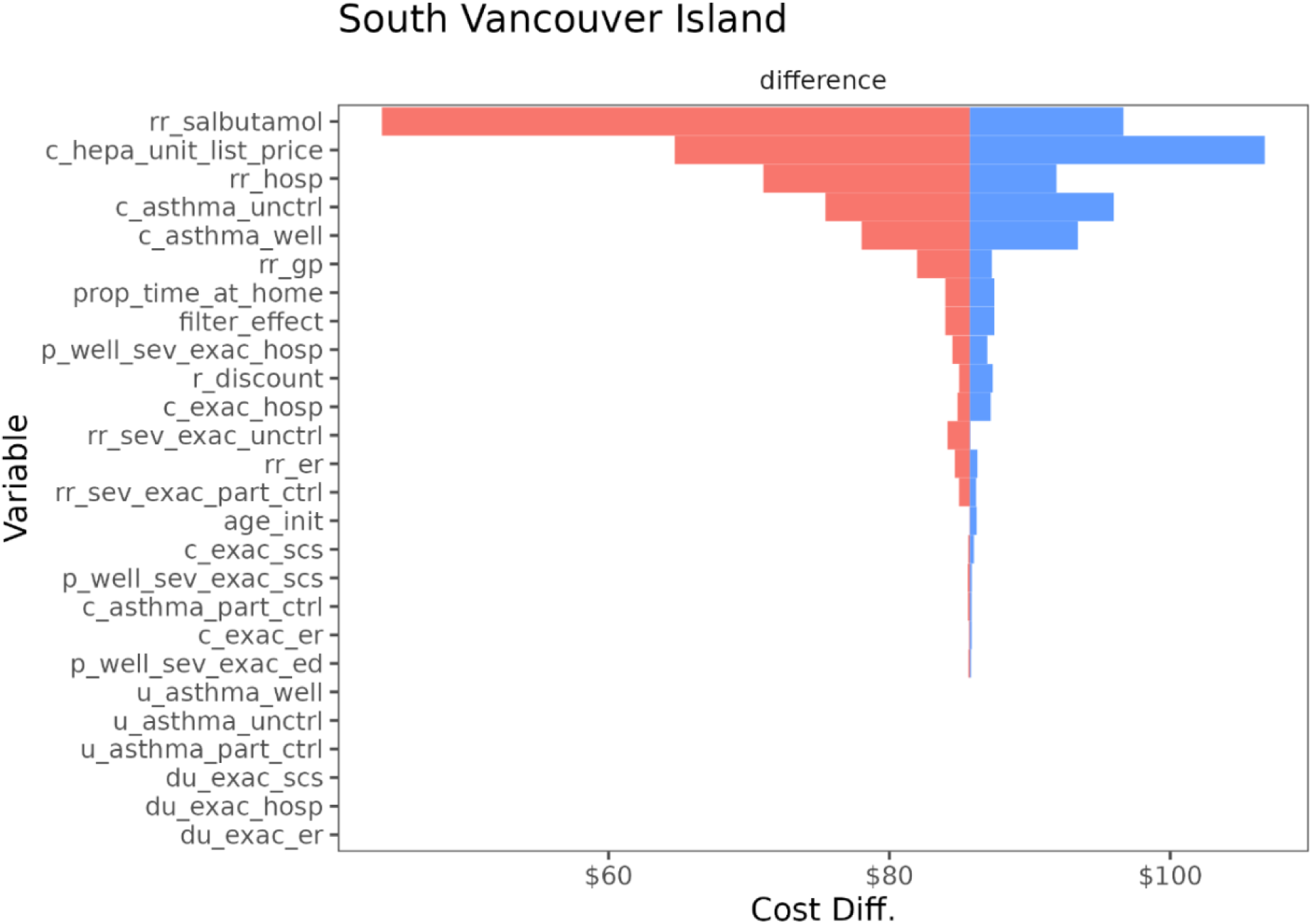
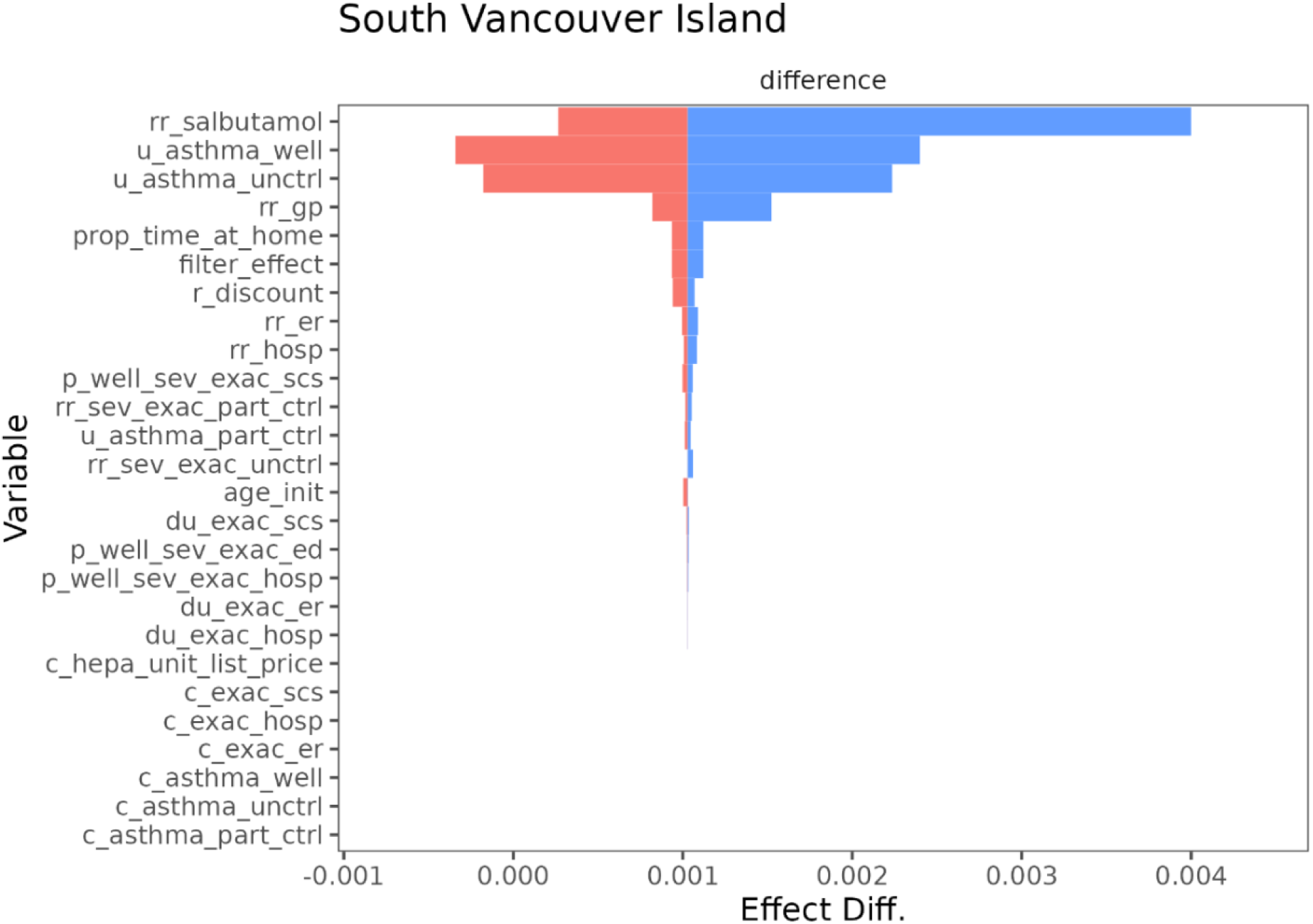
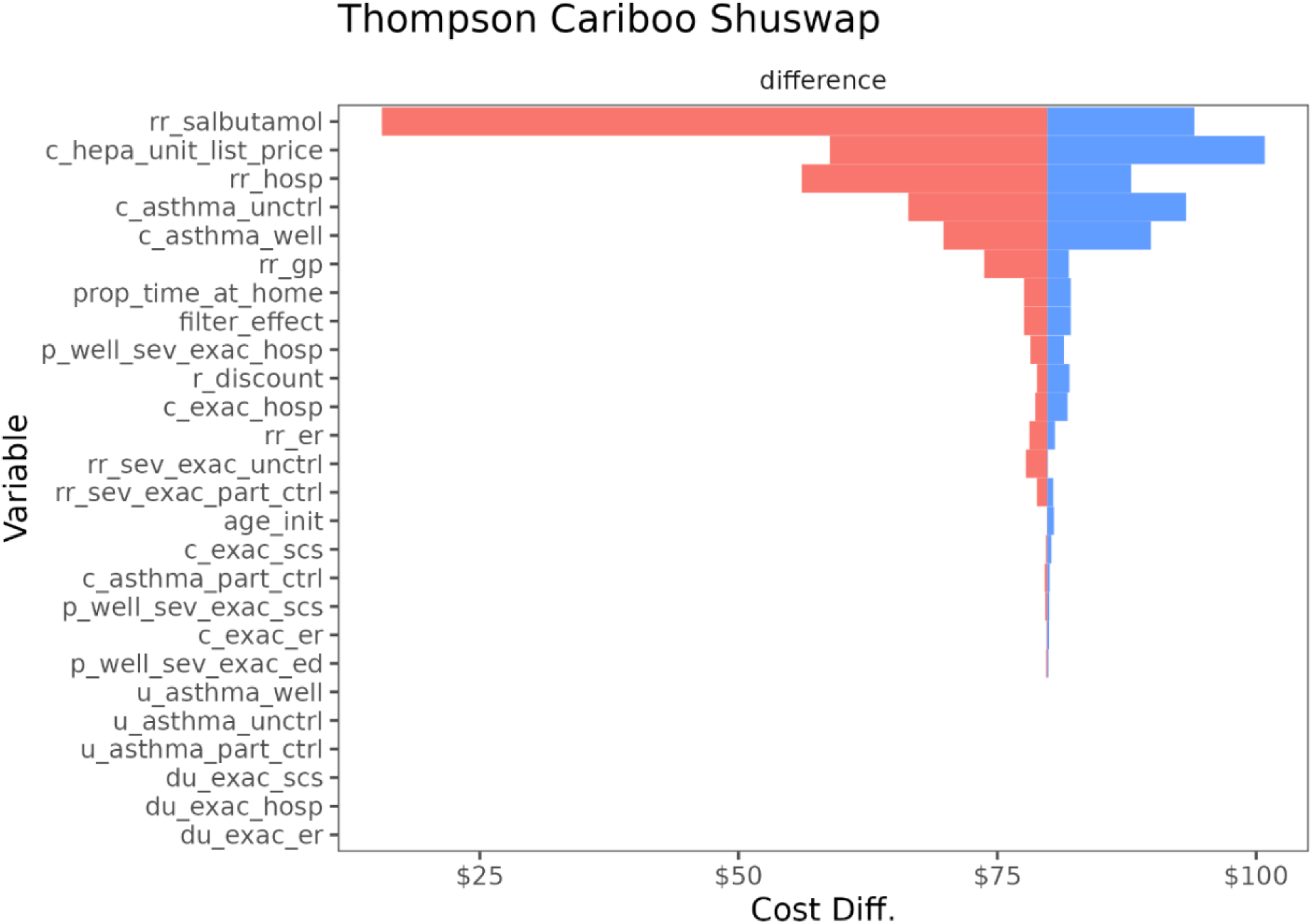
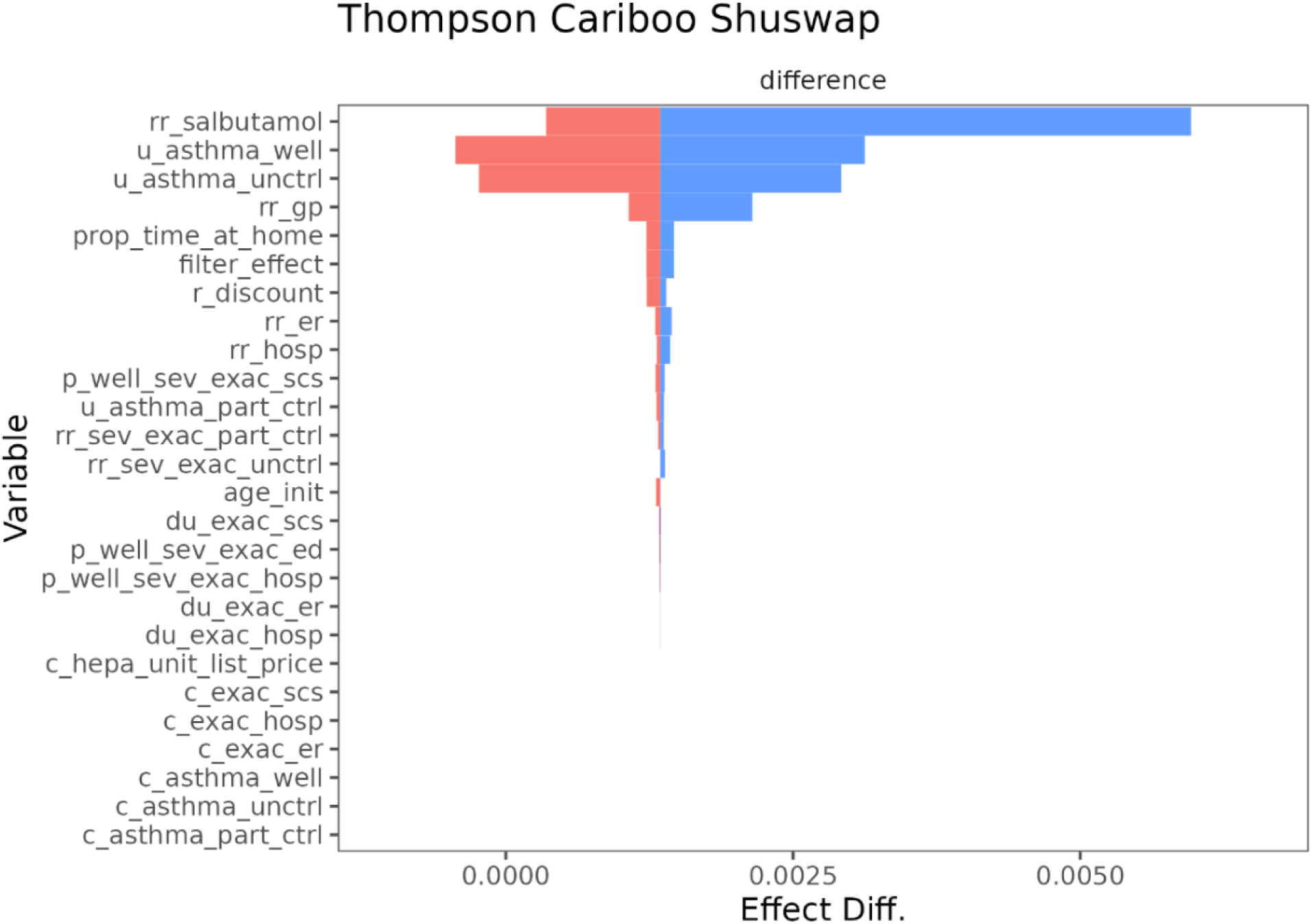
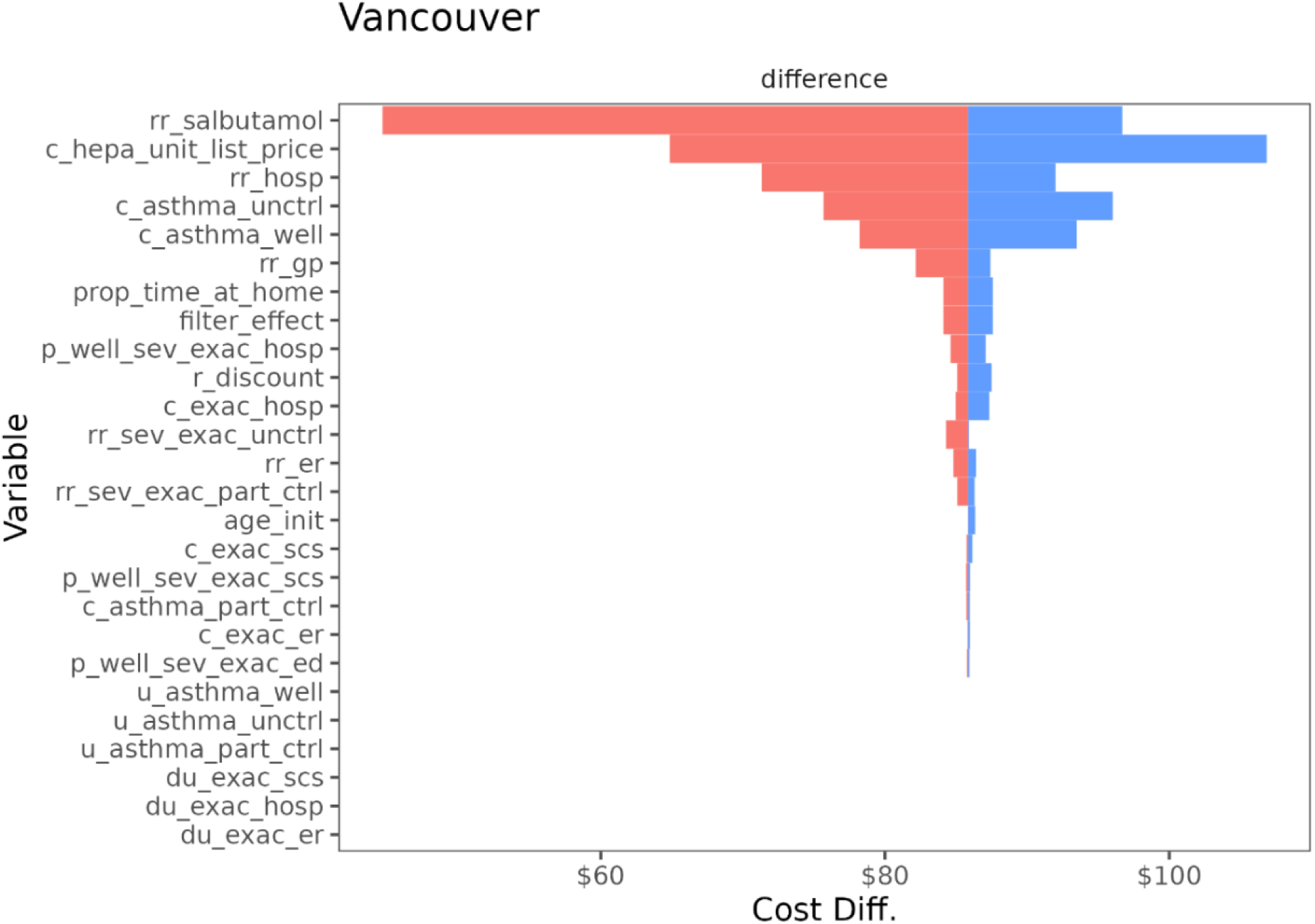
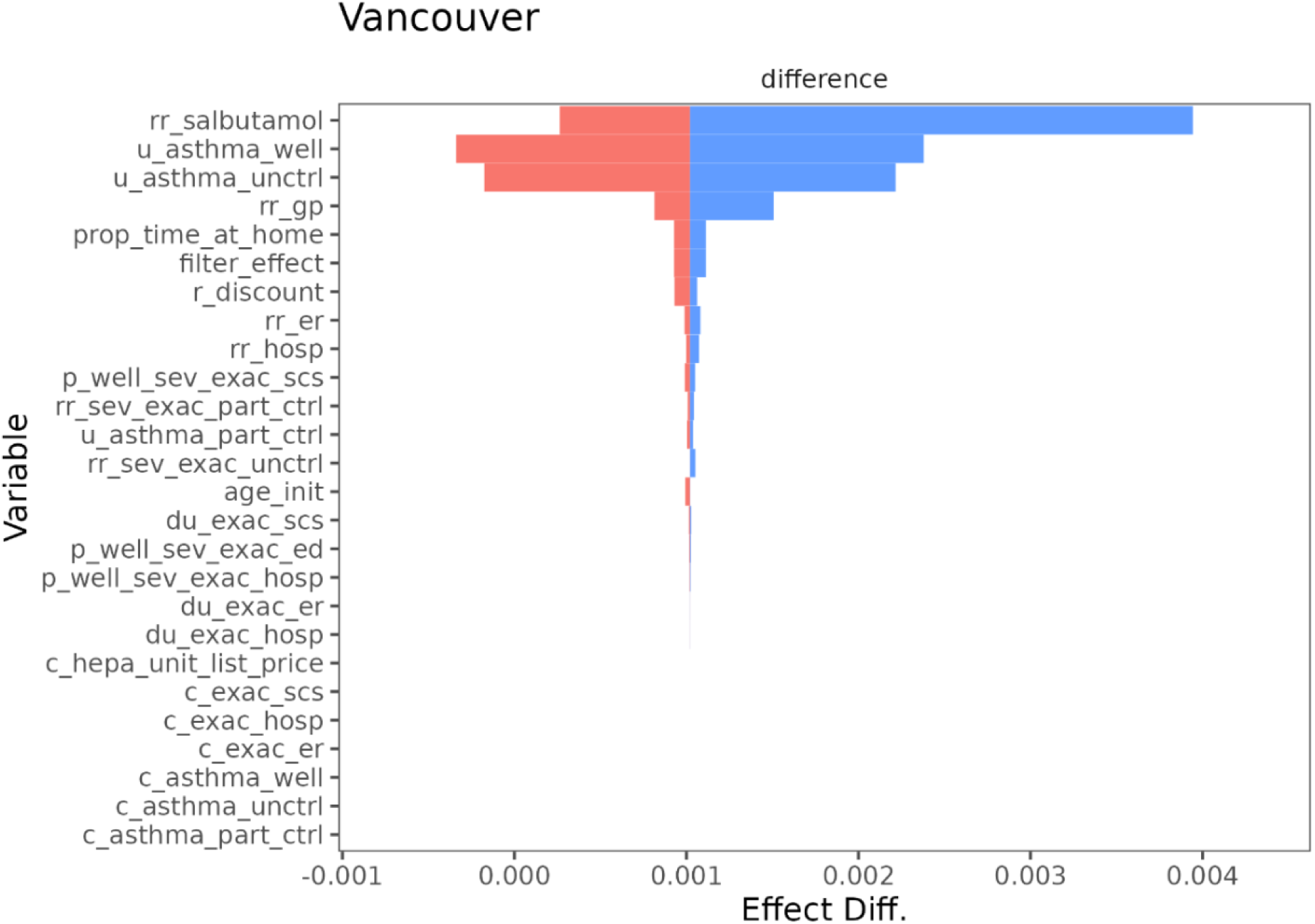

### Appendix: Justifications for Model Parameters

**Table A2:**
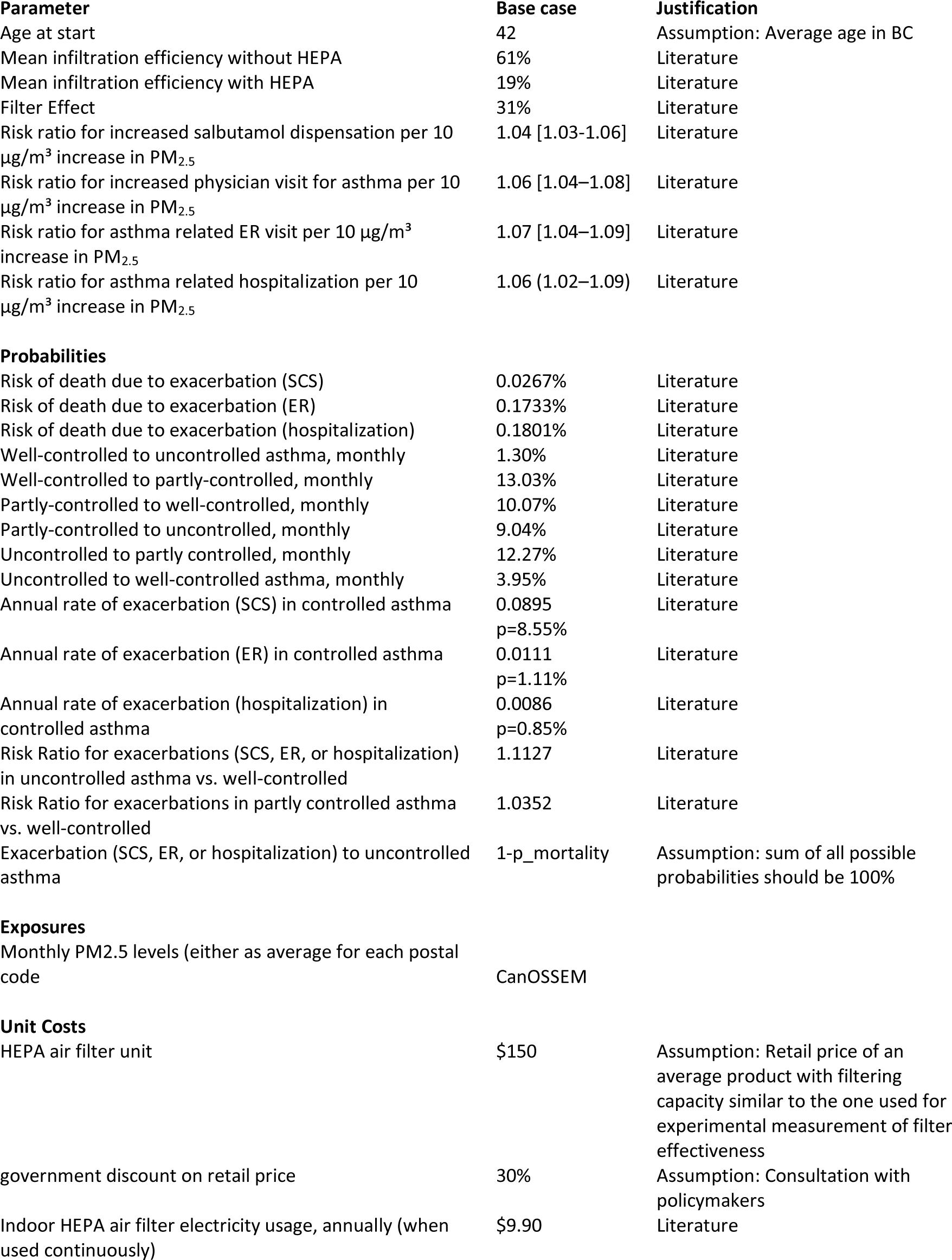
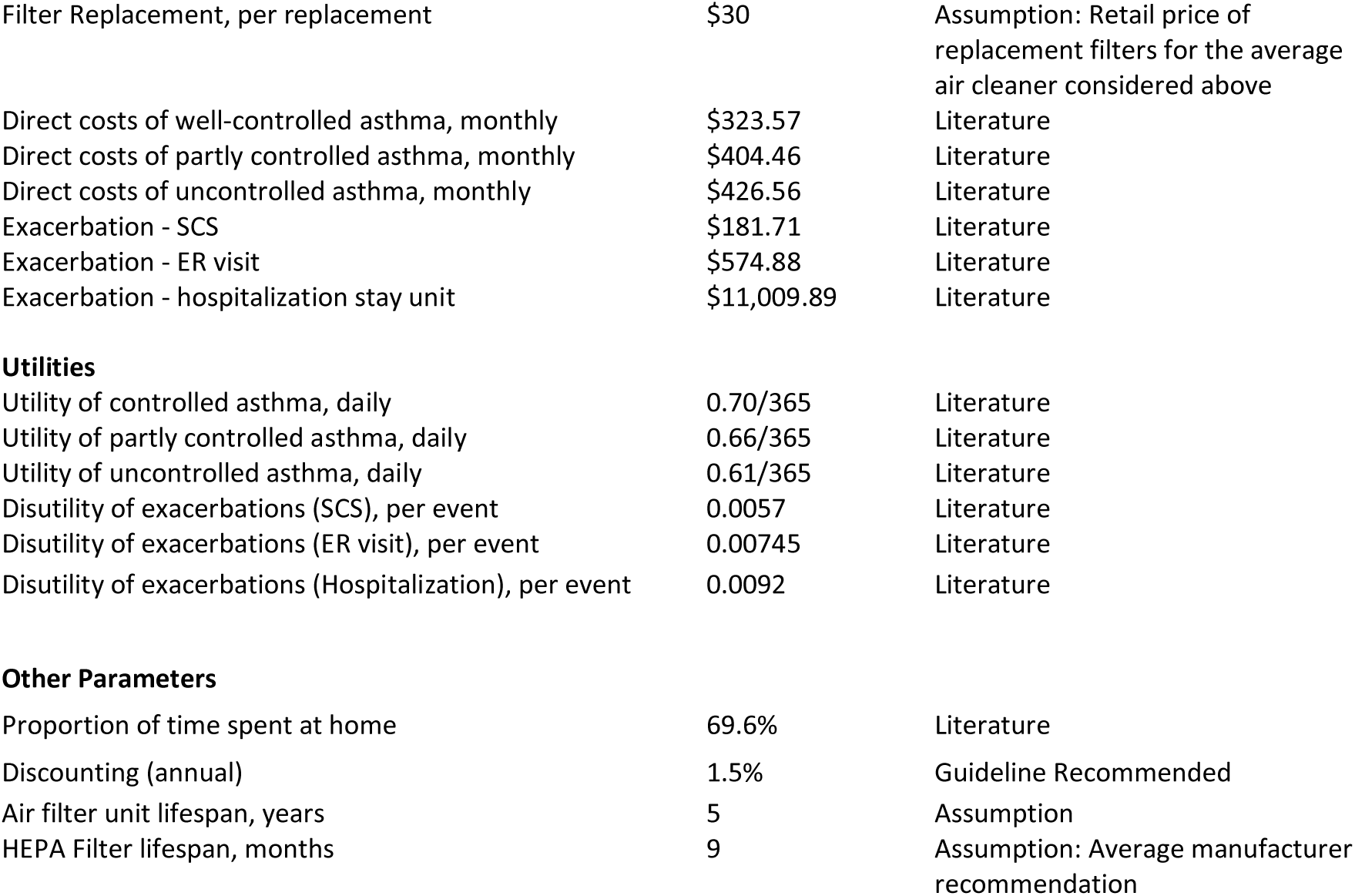
Justifications Model Parameters.

